# Optimizing Antidepressant Efficacy: Generalizable Multimodal Neuroimaging Biomarkers for Prediction of Treatment Response

**DOI:** 10.1101/2024.04.11.24305583

**Authors:** Xiaoyu Tong, Kanhao Zhao, Gregory A. Fonzo, Hua Xie, Nancy B. Carlisle, Corey J. Keller, Desmond J. Oathes, Yvette Sheline, Charles B. Nemeroff, Madhukar Trivedi, Amit Etkin, Yu Zhang

## Abstract

Major depressive disorder (MDD) is a common and often severe condition that profoundly diminishes quality of life for individuals across ages and demographic groups. Unfortunately, current antidepressant and psychotherapeutic treatments exhibit limited efficacy and unsatisfactory response rates in a substantial number of patients. The development of effective therapies for MDD is hindered by the insufficiently understood heterogeneity within the disorder and its elusive underlying mechanisms. To address these challenges, we present a target-oriented multimodal fusion framework that robustly predicts antidepressant response by integrating structural and functional connectivity data (sertraline: R^2^ = 0.31; placebo: R^2^ = 0.22). Remarkably, the sertraline response biomarker is further tested on an independent escitalopram-medicated cohort of MDD patients, validating its generalizability (p = 0.01) and suggesting an overlap of psychopharmacological mechanisms across selective serotonin reuptake inhibitors. Through the model, we identify multimodal neuroimaging biomarkers of antidepressant response and observe that sertraline and placebo show distinct predictive patterns. We further decompose the overall predictive patterns into constitutive *network constellations* with generalizable structural-functional co-variation, which exhibit treatment-specific association with personality traits and behavioral/cognitive task performance. Our innovative and interpretable multimodal framework provides novel and reliable insights into the intricate neuropsychopharmacology of antidepressant treatment, paving the way for advances in precision medicine and development of more targeted antidepressant therapeutics.

**Trial Registration:** Establishing Moderators and Biosignatures of Antidepressant Response for Clinical Care for Depression (EMBARC), NCT#01407094

## Introduction

Major depressive disorder (MDD) is a pervasive condition that profoundly diminishes quality of life for individuals across ages and demographic groups. Unfortunately, current depression treatments are marked by limited effectiveness and unsatisfactory response rates. The development of effective treatments for MDD is hindered by its insufficiently understood heterogeneity and elusive pathophysiology. Encouragingly, over the past decade, neuroimaging studies have successfully identified promising biomarkers for diagnosing MDD^1, 2^, delineating symptom profiles^3^, and predicting treatment responsiveness^4–9^. Recent research has further delved into individual-level antidepressant response^10, 11^, shedding light on the heterogeneity among MDD patients. With the ability to measure physiological neural activity and potentially apply these measures to inform treatment of real patients, neuroimaging studies hold great potential in translating knowledge into precision medicine, thus advancing clinical practice for MDD patients.

Nonetheless, two major challenges persist. First, a substantial proportion of MDD patients experience relapse following discontinuation of an initial successful treatment^12–14^, suggesting that antidepressant medications primarily induce transient neural changes. This raises an intriguing question: how do these transient neural alterations become long-lasting to enable sustained remission? Notably, recent advances in antidepressant response biomarkers are largely based on functional connectivity (FC)^10, 11^, which captures brain region connectivity but may not effectively reflect the enduringness of connections^15^. Alternatively, structural connectivity (SC) provides information about the anatomical basis of neural connections that is complementary to the regional synchronization information provided by FC. Therefore, leveraging a multimodal framework to integrate structural and functional information is essential. While numerous recent studies have pioneered multimodal fusion techniques in an unsupervised manner^16–19^, the resultant fused features are task-agnostic, thus may contain out-of-interest information that potentially leads to overfitting and compromises interpretability. In contrast, a supervised multimodal fusion process guided by prediction targets holds significant potential for improving prediction performance and interpretability. Moreover, the complicated mechanisms of action underlying antidepressants remain poorly understood. Comprehending MDD’s intricate psychopharmacology requires dissecting the overall predictive pattern into constitutive components. Furthermore, the placebo effect in antidepressant response remains inadequately explored^20, 21^. A recent study has suggested a significant contribution of the placebo effect to rapid antidepressant effect^22^. Therefore, caution is warranted when interpreting antidepressant biomarkers, and further research is necessary to identify characteristics that differentiate placebo and drug effects.

To address these challenges, we have developed an innovative target-<u>o</u>riented <u>m</u>ulti<u>m</u>odal fusion (TOMMF) framework, which extracts treatment-specific latent space neuroimaging features by integrating SC and FC data to predict antidepressant response (Fig. 1a). By combining SC and FC, our objective is to identify comprehensive pre-treatment biomarkers to achieve MDD patient stratification with more predictable antidepressant response. Importantly, by integrating the multimodal fusion with prediction task, we introduced guidance from target on latent features, achieving the multimodal fusion in a supervised way that improved prediction performance. By further incorporating the L0-regularization technique^23^, which discourages the contribution of a single feature to multiple dimensions thus resulting in pseudo-exclusive dimension compositions, our approach has the ability in isolating distinct and highly interpretable components within antidepressant response biomarkers (Fig. 1b). Leveraging these distinct components in the multimodal biomarkers for antidepressant response, which we named as *network constellations* (Fig. 1c), we further investigate the characteristic SC-FC co-variation in MDD patients, along with their treatment-specific associations with personality traits and behavioral/cognitive task performance. Our study aims to unravel the distinct roles of structural and functional neural connectivity in predicting antidepressant response, their interplay, and how their treatment-specific patterns can differentiate placebo and drug effects. Importantly, we also assess the generalizability of biomarker findings on an independent cohort. These efforts aim to offer reliable novel insights into the intricate neuropsychopharmacology of antidepressant treatment and lays the groundwork for improved clinical practice in MDD treatment.

**Fig. 1.**
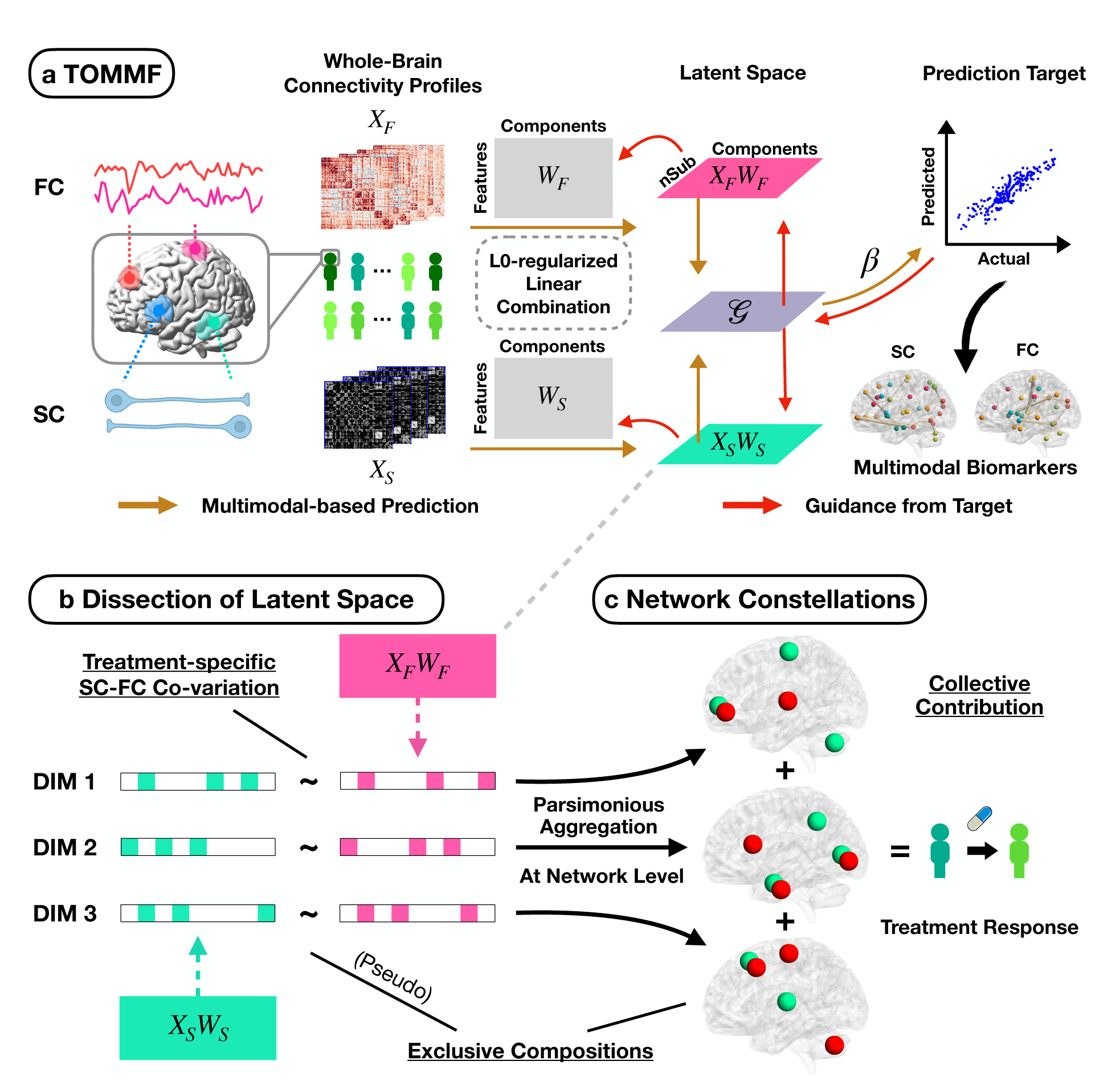
<u>T</u>arget-<u>o</u>riented <u>m</u>ulti<u>m</u>odal fusion (TOMMF) framework. **a** The overall framework. Structural and functional connectivity (SC and FC) features are extracted from structural and resting-state functional MRI. Subsequently, the framework optimizes coefficients of multimodal fusion (W_S_ and W_F_) to transform the SC and FC features (X_S_ and X_F_) into a latent space 𝓖 that distills the co-variation in SC and FC. Importantly, 𝓖 may lean towards a particular modality given its relative importance. Meanwhile, the framework optimizes the coefficients (***β***) of latent features to predict the target (e.g., antidepressant response). Notably, in this framework with bidirectional propagation, the target not only benefits from the multimodal-derived latent features, but also informs their calculation. The latent space 𝓖 essentially relays the information between the prediction target and each of the data modalities, thus integrating the information from SC, FC, and the target. L0-regularization is imposed to control model complexity and facilitates the identification of distinct latent dimensions. Guidance from target is confined to the training set to prevent information leakage. Multimodal biomarkers for the target are derived from coefficients in cross-validated TOMMF-based prediction models. **b** Dissection of latent space. The overall latent space is essentially a compilation of dimensions, with each dimension incorporating a unique treatment-specific combination of associated SC and FC features. The L0-regularization applied on W_S_, W_F_, and ***β*** discourages the contribution of a single feature to multiple dimensions, resulting in pseudo-exclusive dimension compositions. **c** Network constellations. Network constellations are constructed by parsimoniously aggregating SC and FC features with the strongest correlations at the network level. These constellations allocate network-level connectivity features in an exclusive manner, enhancing the clarity of the resulting psychopharmacological pathways. Importantly, these network constellations (as representations of latent dimensions) contribute to the treatment response prediction in a collective manner.

## Results

### Multimodal Neuroimaging Biomarkers for Sertraline Response

We first applied the TOMMF framework to 103 MDD patients receiving sertraline treatment from the EMBARC dataset^24^, developing a prediction model for individual antidepressant response based on SC and FC extracted for 100 cortical^25^ and 35 subcortical^26, 27^ regions-of-interest (ROIs). Antidepressant response was quantified as the pre- to post-treatment change in 17-item Hamilton Depression Rating Scale scores. The model exhibited strong cross-validation (10x ten-fold) performance in predicting sertraline response (R^2^ = 0.3149, r = 0.5689, p = 3.63 x 10^-10^, Fig. 2a), with its significance confirmed by permutation testing (p_perm_ < 0.001, SFig. 1a). Notably, the sertraline response prediction model demonstrated no predictability to placebo response (R^2^ < 0, r = -0.0312, p = 0.7352, Fig. 2b), suggesting its specificity to sertraline. Furthermore, the TOMMF framework outperformed various baseline methods, including regression models trained with concatenated SC and FC features, sparse canonical correlation analysis-based fused features, partial least squares regression, and single data modalities (one-tailed Wilcoxon signed-rank test: all p_FDR_ ≤ 0.0020, Fig. 2c). Collectively, these findings underscore the potential of the TOMMF framework in providing enhanced predictive power for sertraline response on the individual patient level and identifying neural phenotypic characteristics that can be exploited for treatment stratification.

**Fig. 2.**
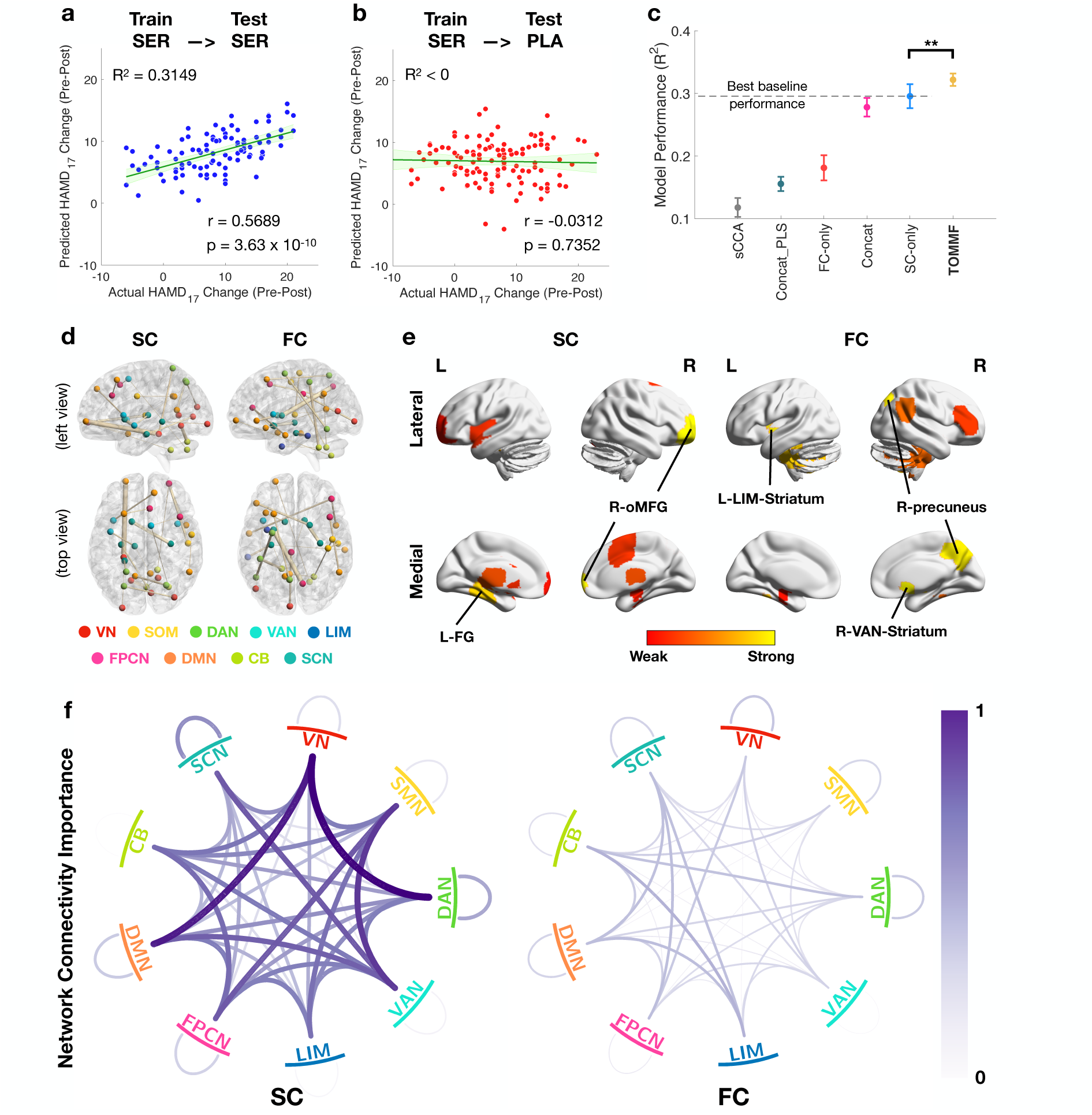
Multimodal neuroimaging biomarkers for sertraline-induced antidepressant response. **a** Multimodal-based sertraline response prediction model. The model demonstrated robust prediction performance for sertraline response evaluated by 10x 10-fold cross-validation. **b** Specificity of sertraline response prediction model to treatment. The model for sertraline response showed no prediction capability for placebo response. **c** Comparison of target-<u>o</u>riented <u>m</u>ulti<u>m</u>odal <u>fusion</u> (TOMMF) framework with baseline methods. The TOMMF framework showed significantly higher prediction performance than baseline methods, including partial least squares regression with concatenated SC and FC features (Concat_PLS), LASSO regression with only SC, only FC, sparse canonical correlation analysis-based fused features (sCCA), or concatenated SC and FC features (Concat). The statistical significance is evaluated by one-tailed paired t-test (p = 0.0024) with 100x 10-fold cross-validation. The error bars indicate the standard error. **d** Important FC and SC features for sertraline response. The predictive pattern exhibited a multivariate foundation encompassing every functional module of the brain. Specifically, the connection between left fusiform gyrus and left middle frontal gyrus showed the strongest contribution among structural connectivity, while the connection between right precuneus and left amygdala displayed the strongest contribution among functional connectivity. The top 20 connectivity features are shown for each data modality for clarity. **e** Important brain regions in the multimodal predictive pattern for sertraline response. The right orbital part of the middle frontal gyrus (R-oMFG) and the left fusiform gyrus (L-FG) showed most substantial contribution in abnormal structural connectivity. The bilateral striata and the right precuneus played crucial roles in abnormal functional connectivity. Within the left striatum, the region with rich connections to the limbic network (L-LIM-Striatum) demonstrated highest importance. Within the right striatum, the region functionally associated with the ventral attention network (R-VAN-Striatum) held particular importance. For visualization purpose, the striata are projected to the nearest surface. The top 10 brain regions are shown for each data modality for clarity. **f** Multimodal network connectivity importance for sertraline response prediction. The network connectivity importance is calculated as the average absolute value of non-zero ROI-level connectivity weights of the prediction model. Overall, the SC features show greater importance than FC features. The VN-DAN and VN-DMN SCs make the strongest contribution to sertraline response prediction. **VN**: Visual Network. **SOM**: Somatomotor Network. **DAN**: Dorsal Attention Network. **VAN**: Ventral Attention Network. **LIM**: Cortical Limbic Network. **FPCN**: Fronto-Parietal Control Network. **DMN**: Default-Mode Network. **CB**: Cerebellum. **SCN**: Subcortical Network.

Afterward, we examined the multimodal neuroimaging biomarkers for sertraline response as dictated by the prediction model weights. Overall, the predictive pattern exhibited a robust multivariate foundation encompassing SC and FC involving every functional brain network (Fig. 2d). Specifically, the connection between left fusiform gyrus and left middle frontal gyrus displayed the most substantial contribution among SC, while the connection between right precuneus and left amygdala demonstrated the strongest contribution among FC. Further investigation of pivotal brain regions in the structural and functional profiles of sertraline response biomarkers (Fig. 2e) revealed that the right orbital part of the middle frontal gyrus and the left fusiform gyrus played key roles in the SC-based biomarker, and the bilateral striata and the right precuneus played were prominent in the FC-based biomarker. Notably, within the left striatum, the region functionally associated with the ventral attention network held particular importance, while the region with rich connections to the limbic network played a crucial role in the right striatum. Lastly but importantly, we examined the network connectivity importance by averaging the absolute non-zero weights of ROI-level connectivity features. As a result, we found that SC exhibited overall greater importance than FC, while the SC between visual network (VN) and dorsal attention network (DAN) and between VN and default mode network (DMN) displayed the strongest contribution to sertraline response prediction (Fig. 2f).

### Multimodal Neuroimaging Biomarkers for Placebo Response

Subsequently, we applied the TOMMF framework to derive a prediction model for predicting placebo response in 120 placebo-medicated MDD patients. As a result, the framework exhibited high cross-validation performance for placebo response prediction (R^2^ = 0.2190, r = 0.4682, p = 6.96 x 10^-8^, Fig. 3a), whose significance was further confirmed by permutation test (p_perm_ = 0.001, SFig. 1b). Importantly, the placebo response prediction model was unable to predict sertraline response (R^2^ < 0, r = -0.0597, p = 0.5492, Fig. 3b), suggesting its specificity to placebo. Furthermore, our target-oriented multimodal framework showed superiority of placebo response prediction over baseline methods (one-tailed Wilcoxon signed-rank test: all p_FDR_ ≤ 9.77 x 10^-4^, Fig. 3c). Together, these results demonstrated the potential of this framework in providing enhanced predictive power for placebo response and showcased its generalizability to different clinical prediction targets.

**Fig. 3.**
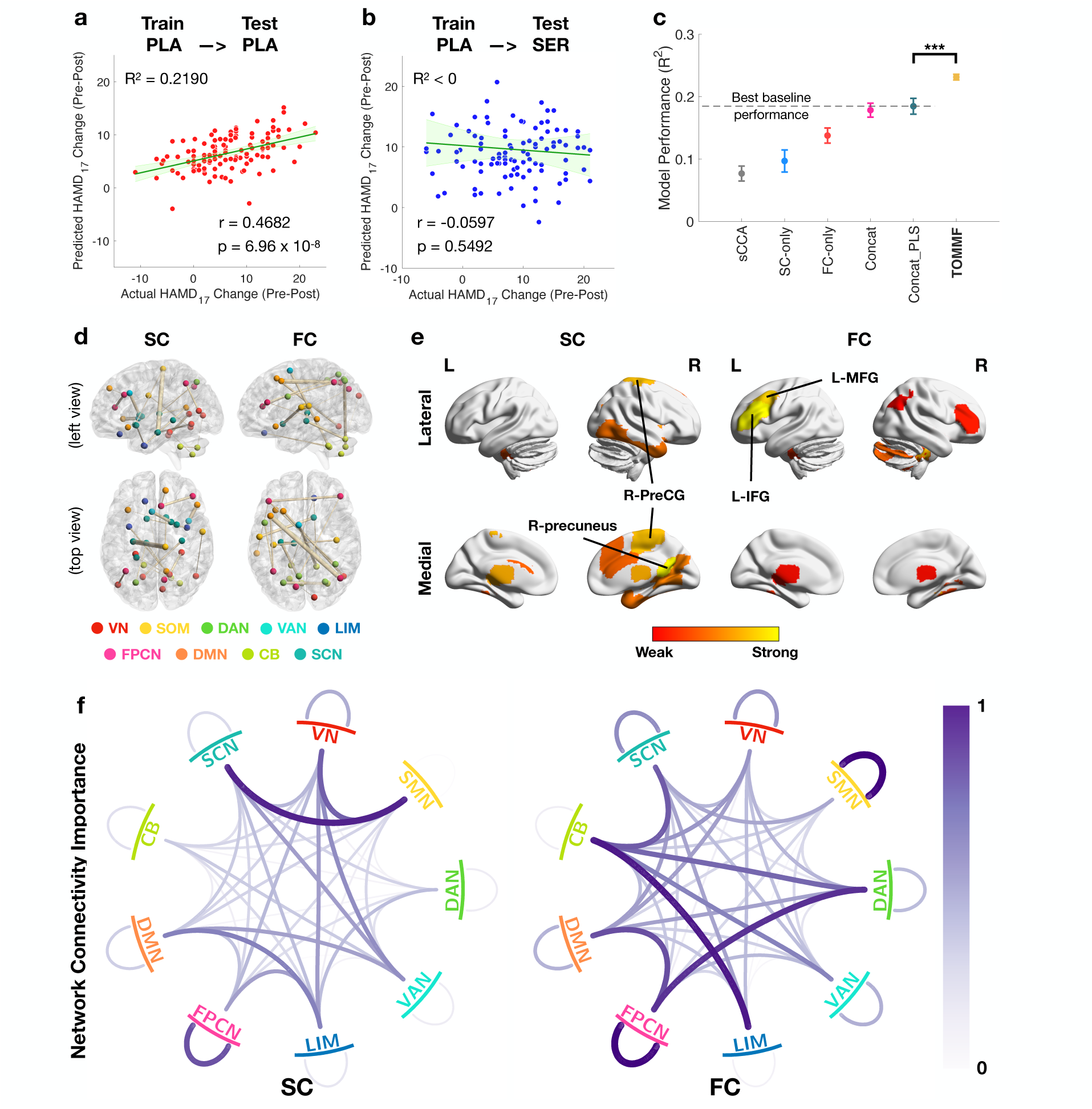
Multimodal neuroimaging biomarkers for placebo-induced antidepressant response. **a** Multimodal-based placebo response prediction model. The model demonstrated robust prediction performance for placebo response evaluated by 10x 10-fold cross-validation. **b** Specificity of placebo response prediction model to treatment. The model for placebo response showed no prediction capability for sertraline response. **c** Comparison of TOMMF framework with baseline methods. The TOMMF framework showed significantly higher prediction performance than baseline methods, including partial least squares regression with concatenated SC and FC features (Concat_PLS), LASSO regression with only SC, only FC, sparse canonical correlation analysis-based fused features (sCCA), or concatenated SC and FC features (Concat). The statistical significance is evaluated by one-tailed paired t-test (p = 9.74 x 10^-6^) with 100x 10-fold cross-validation. The error bars indicate the standard error. **d** Important FC and SC features for placebo response. The predictive pattern exhibited a multivariate foundation encompassing every functional module of the brain. Specifically, the connection between right precentral gyrus (R-PreCG) and left anterior hippocampus showed the strongest contribution among structural connectivity, while the connection between left middle frontal gyrus (L-MFG) displayed the strongest contribution among functional connectivity. The top 20 connectivity features are shown for each data modality for clarity. **e** Important brain regions in the multimodal predictive pattern for placebo response. The right precuneus and the R-PreCG showed most substantial contribution in abnormal structural connectivity. The left inferior frontal gyrus (L-IFG) and the L-MFG played crucial roles in abnormal functional connectivity. The top 10 brain regions are shown for each data modality for clarity. **f** Multimodal network connectivity importance for placebo response prediction. The network connectivity importance is calculated as the average absolute value of non-zero ROI-level connectivity weights of the prediction model. Overall, the FC features show greater importance than SC features. The within-FPCN, within-SMN FCs, and SMN-SCN SC make the strongest contribution to placebo response prediction. **VN**: Visual Network. **SOM**: Somatomotor Network. **DAN**: Dorsal Attention Network. **VAN**: Ventral Attention Network. **LIM**: Cortical Limbic Network. **FPCN**: Fronto-Parietal Control Network. **DMN**: Default-Mode Network. **CB**: Cerebellum. **SCN**: Subcortical Network.

We then explored the multimodal neuroimaging biomarkers for placebo response as guided by the prediction model weights. Similar to the biomarkers for sertraline response, the predictive pattern for placebo response exhibited a robust multivariate foundation encompassing every functional brain network (Fig. 3d). Specifically, the connection between right precentral gyrus and left anterior hippocampus displayed the most substantial contribution among SC, while the connection between left middle frontal gyrus and right angular gyrus demonstrated the strongest contribution among FC. Further investigation of important brain regions in the structural and functional profiles of placebo response biomarkers (Fig. 3e) revealed that the right precuneus and the right precentral gyrus played key roles in the SC-based biomarker, while the left inferior frontal gyrus and the left middle frontal gyrus were prominent in the FC-based biomarker. Network connectivity importance revealed that FC provided overall greater contribution for placebo response than SC, while the FCs within somatomotor network (SMN) and within frontoparietal control network (FPCN) and the SC between SMN and subcortical network (SCN) showed particular importance (Fig. 3f).

### Generalizability of Treatment Response Biomarkers on Independent Cohort

We then examined the generalizability of treatment response biomarkers on an independent cohort. Remarkably, despite the differences in medication and MDD symptom assessment (see **Methods** for details), the symptom relief predicted by sertraline response biomarker exhibited a significant correlation with the escitalopram-induced one in the CAN-BIND-1 cohort (r = 0.2739, p = 0.0104, Fig. 4a). This result demonstrated the generalizability of the multimodal sertraline response biomarker and suggested a potential overlap of psychopharmacological mechanisms across selective serotonin reuptake inhibitors. In contrast, no such correlation was observed between the escitalopram-induced symptom relief and the one predicted by placebo response biomarker (r = -0.0499, p = 0.6603, Fig. 4b), consistent with our finding that sertraline and placebo alleviate the symptoms via distinct mechanisms.

**Fig. 4.**
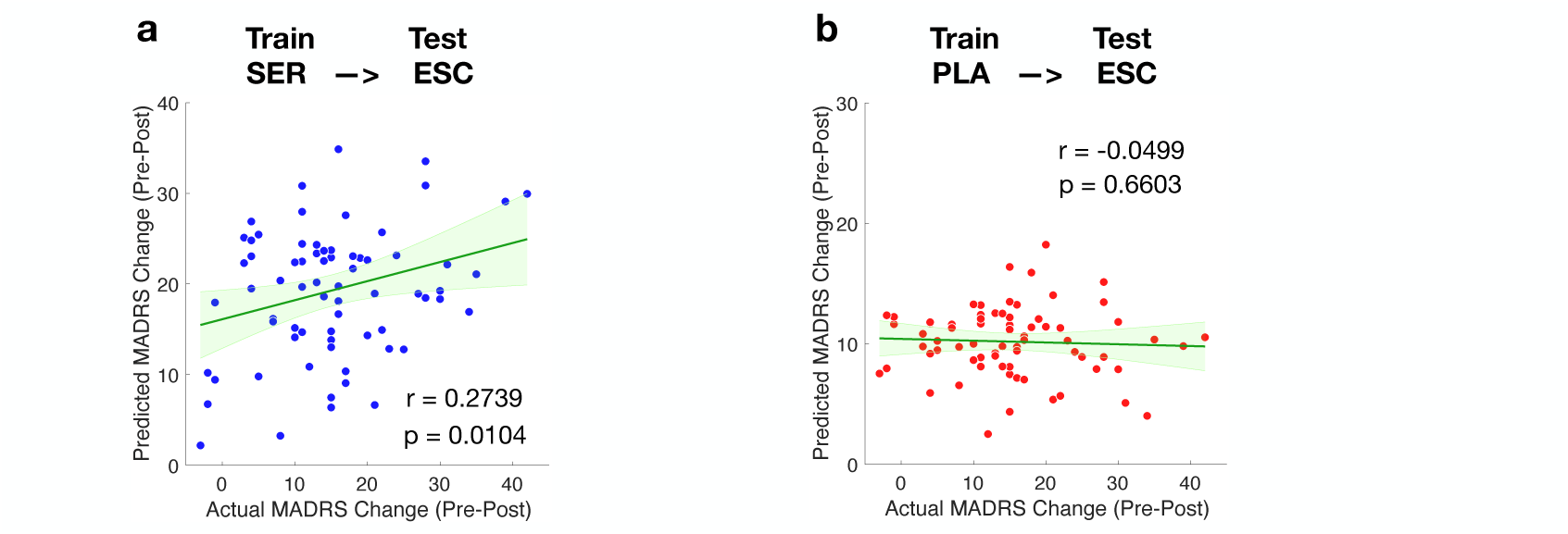
Generalization analysis of multimodal biomarkers for sertraline and placebo response in the CAN-BIND-1 cohort (n = 71). Multimodal-based prediction models for sertraline (**a**) and placebo (**b**) response were applied to an independent cohort of escitalopram-medicated MDD patients, with symptom relief assessed by the Montgomery–Åsberg Depression Rating Scale (MADRS). Predicted HAMD_17_ changes were transformed to MADRS changes using an equipercentile-based conversion (MADRS ≈ HAMD_17_ x 1.3). Despite differences in medication, symptom relief predicted by the sertraline response model significantly correlated with actual escitalopram response, validating the generalizability of the sertraline response biomarker and suggesting an overlap of pharmacological mechanisms across selective serotonin reuptake inhibitors. In contrast, the placebo response model did not predict escitalopram response, consistent with our findings that sertraline and placebo alleviate the symptoms via distinct mechanisms. P-values are one-tailed as we seek only positive correlations.

### Distinct Data Modality Importance for Sertraline and Placebo

After validating the identified biomarkers, we investigated how SC and FC contribute to the sertraline and placebo response respectively. To this end, we examined the effect of the modality importance parameter *ρ_Λ_* on prediction performance (see **Methods** for details). Interestingly, our findings indicated that the sertraline response biomarker exhibited a preference for SC, while the placebo response biomarker showcased a more substantial influence of FC (SFig. 2). These results suggested the distinct biomarkers for sertraline and placebo responses, implying the potential difference in their psychopharmacological mechanisms.

### Network Constellations with Generalizable SC-FC Co-variation for MDD patients

To further understand the essence of latent features and parse the intricate psychopharmacology of MDD, we constructed network constellations by aggregating network components with the highest cross-modality correlations based on the sertraline response biomarker (see **Methods** for details). Essentially, we aimed to identify co-variations of SC and FC that are predictive to antidepressant response. As a result, we identified three network constellations with generalizable SC-FC co-variation spanning the intrinsic-extrinsic functioning axis. We named these network constellations based on the pattern of findings, including the default-mode regulatory constellation, the affective constellation, and the sensory processing constellation (Fig. 5a). The default-mode regulatory constellation consisted of the intra-connectivity within DMN, SCN, and their connectivity with DAN and FPCN. The affective constellation was primarily composed of the intra-connectivity of cortical limbic network (LIM) and its connectivity with VN, SMN, and DAN. The sensory processing constellation encompassed the VN’s connectivity to DMN and SCN, as well as connectivity involving attention networks and cerebellum. For the sertraline response biomarker, the sertraline-medicated patients indeed showed significant correlations between the SC and FC constellation scores (default-mode regulatory: r = 0.4385, p = 1.96 x 10^-12^; affective: 0.3027, p = 1.62 x 10^-6^; sensory processing: r = 0.1375, r = 0.0190), which were successfully generalized to the placebo-medicated patients (default-mode regulatory: r = 0.1394, p = 0.0154; affective: r = 0.1579, p = 0.0072; sensory processing: 0.1445, p = 0.0126, top row of Fig. 5b).

**Fig. 5.**
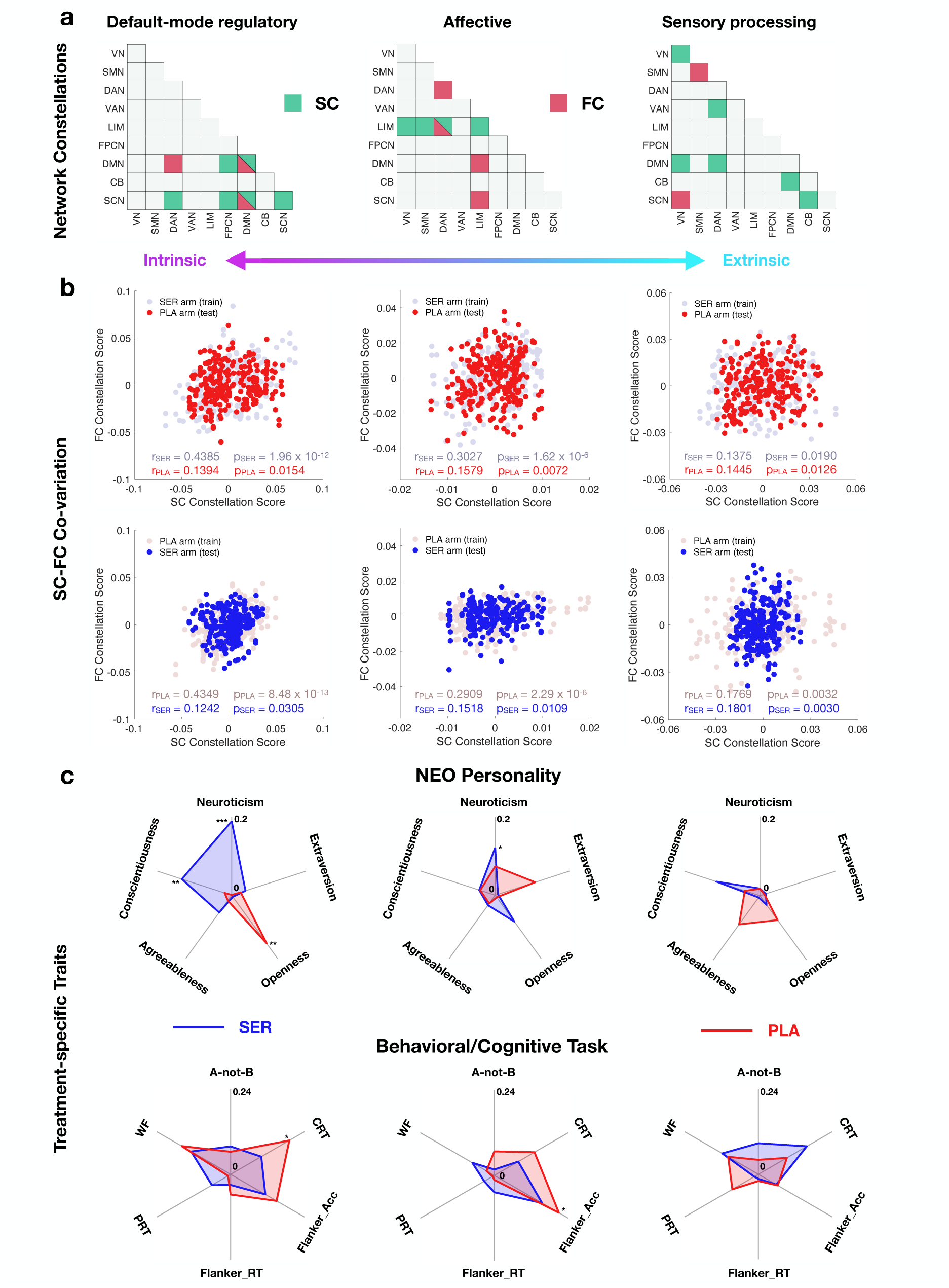
Network constellations in multimodal biomarkers for sertraline and placebo response. **a** Composition of network constellations. First, network components are quantified as the sum of region-level connectivity scores involving a particular network pair, weighted by antidepressant response model coefficients. Subsequently, network constellations are constructed by aggregating network components with the highest cross-modality correlations. Three generalizable network constellations spanning the intrinsic-extrinsic functioning axis are identified, including the default-mode regulatory constellation, the affective constellation, and the sensory processing constellation. Notably, SC and FC contributions are not equivalent. **VN**: Visual Network. **SOM**: Somatomotor Network. **DAN**: Dorsal Attention Network. **VAN**: Ventral Attention Network. **LIM**: Cortical Limbic Network. **FPCN**: Fronto-Parietal Control Network. **DMN**: Default-Mode Network. **CB**: Cerebellum. **SCN**: Subcortical Network. **b** Generalizable SC-FC co-variation of MDD patients in network constellations. Top row: SC-FC co-variation of sertraline response biomarker. The SC and FC contributions of constellation scores exhibit significant positive correlation on the sertraline arm, demonstrating the framework have successfully identified the link between SC and FC. As SC-FC co-variation is an intrinsic property of pre-treatment neuroimaging data, we expect observe the the same relationship in the other treatment arm. Encouragingly, the patients receiving placebo indeed show significant positive correlation between SC and FC contributions of all three constellation scores, suggesting the generalizability of SC-FC co-variation of sertraline response. Bottom row: SC-FC co-variation of placebo response biomarker. The network constellations constructed on sertraline response biomarker also show robust SC-FC co-variation with placebo response biomarker for all three network constellations. This SC-FC co-variation is generalizable across treatment arms. **c** Treatment-specific traits of network constellations. Top row: Association of treatment-specific network constellations with NEO personality traits. For default-mode regulatory constellation, the sertraline-specific biomarker exhibits a robust association with neuroticism and conscientiousness, while the placebo-specific biomarker significantly associates with openness. The sertraline-specific biomarker in affective constellation also exhibits significant association with neuroticism, while no biomarkers in sensory processing constellation showed significant association with NEO personality traits. Bottom row: Association of treatment-specific network constellations with behavioral/cognitive task performance. The placebo-specific biomarker in default-mode regulatory constellation shows a significant association with CRT, and the affective constellation score shows significant association with Flanker_Acc. No sertraline-specific constellations exhibit significant correlations with the task performance we have investigated. **A-not-B**: Accuracy in A-not-B task. **CRT**: Choice reaction time. **Flanker_Acc**: Flanker inference on accuracy. **Flanker_RT**: Flanker inference on reaction time. **PRT**: Response bias in probabilistic reward task. **WF**: Total number of valid words in word fluency task.

Afterward, we examined whether these network constellations captured SC-FC co-variation in placebo response biomarker. Encouragingly, these network constellations also exhibited generalizable SC-FC co-variation for the placebo response biomarker (<u>placebo-</u> <u>medicated patients</u>: default-mode regulatory: r = 0.4349, p = 8.48 x 10^-13^; affective: r = 0.2909, p = 2.29 x 10^-6^; sensory processing: r = 0.1769, p = 0.0032; <u>sertraline-medicated patients</u>: default-mode regulatory: r = 0.1242, p = 0.0305; affective: r = 0.1518, p = 0.0109; sensory processing: r = 0.1801, p = 0.0030, bottom row of Fig. 5b), demonstrating the generalizability of the SC-FC co-variation within these identified network constellations for MDD patients.

### Treatment-specific Traits of Network Constellations

While the network constellations displayed generalizable SC-FC co-variation between treatment arms, they might each exhibit distinct contributions to clinical improvements. Therefore, we then investigated whether these network constellations showed treatment-specific traits as reflected in antidepressant response biomarkers. Notably, the SC and FC constellation scores were summed together to get a combined score in this analysis.

First, as a recent study demonstrated significant association between personality traits and neuropsychopathology^28^, we examined the correlations between network constellation scores and NEO personality traits of MDD patients (top row of Fig. 5c). Interestingly, in the default-mode regulatory constellation, the sertraline-specific biomarker exhibited a robust association with neuroticism (r = 0.1897, p_FDR_ = 2.12 x 10^-4^) and conscientiousness (r = -0.1343, p_FDR_ = 0.0098), while the placebo-specific biomarker was significantly associated with openness (r = 0.1524, p_FDR_ = 0.0052). The sertraline-specific biomarker in affective constellation also exhibited significant association with neuroticism (r = 0.1216, p_FDR_ = 0.0451), while no biomarkers in sensory processing constellation showed significant association with NEO personality traits.

Subsequently, we explored the association between network constellation scores and behavioral/cognitive task performance at baseline, including the A-not-B task, choice reaction time, the flanker test, the probabilistic reward task, and the word fluency test (bottom row of Fig. 5c). Results revealed that the placebo-specific biomarker in default-mode regulatory constellation was significantly associated with choice reaction time (r = -0.1909, p_FDR_ = 0.0264), while the affective constellation score showed a significant association with Flanker inference on accuracy (r = 0.2097, p_FDR_ = 0.0106). No sertraline-specific constellation scores exhibited significant correlations with the task performance we investigated.

### Enhanced Robustness of Biomarkers Endowed by TOMMF

Lastly, we examined the association between TOMMF-based multimodal biomarkers and SC/FC biomarkers from unimodal baseline methods for both sertraline and placebo responses. Predictive SC exhibited a strong correlation between unimodal- and multimodal-based patterns (r_SER_ = 0.8186, r_PLA_ = 0.8158, SFig. 3), while predictive FC showed a relatively modest correlation (r_SER_ = 0.3401, r_PLA_ = 0.4352, SFig. 3). This highlighted that the TOMMF framework effectively utilized inputs from each data modality for improved prediction. Notably, the pattern correlation for FC aligned with its relative importance for placebo compared to sertraline, and the overall stronger correlation for SC echoed its relative persistence compared to FC.

Subsequently, the robustness of predictive patterns across cross-validation folds was investigated. We respectively examined the intraclass correlation coefficients (ICC) of predictive SC and FC patterns from 10×10 cross-validation folds demonstrated robustness for both sertraline and placebo response in the TOMMF framework (ICC_SC,SER_ = 0.5326, 95% CI [0.5253,0.5400]; ICC_FC,SER_ = 0.3916, 95% CI [0.3846,0.3988]; ICC_SC,PLA_ = 0.4801, 95% CI [0.4727, 0.4875]; ICC_FC,PLA_ = 0.4926, 95% CI [0.4853, 0.5001]). Importantly, these were higher than the robustness of predictive patterns obtained from unimodal methods (ICC_SC,SER_ = 0.4395, 95% CI [0.4322,0.4468]; ICC_FC,SER_ = 0.3018, 95% CI [0.2955,0.3081]; ICC_SC,PLA_ = 0.3972, 95% CI [0.3901,0.4044]; ICC_FC,PLA_ = 0.3169, 95% CI [0.3105, 0.3234]), with statistical significance confirmed by one-tailed two-sample z-tests (p_SC,SER_ = 1.11 x 10^-16^, p_FC,SER_ = 3.09 x 10^-12^, p_SC,PLA_ = 2.41 x 10^-12^, p_FC,PLA_ < 10^-323^). Collectively, these results suggested that the TOMMF framework enhanced the predictability of antidepressant response by integrating information across data modalities and improved the robustness of identified patterns for each data modality.

## Discussion

In this study, we introduced the TOMMF framework to investigate the intricate neuropsychopharmacology of antidepressant treatment. We explored the roles and interplay of structural and functional neural connectivity and their treatment-specific patterns that distinguish placebo and drug effects. The integration of SC and FC facilitated the development of predictive models for sertraline- and placebo-induced antidepressant responses that outperformed unimodal methods. These models revealed critical brain connections/regions linked to antidepressant response and the associations between SC and FC abnormalities. Remarkably, the sertraline response biomarker exhibited significant generalizability on an independent escitalopram-medicated cohort, holding promise for enabling applicable neuroimaging-guided antidepressant treatment and suggesting a shared mechanism across selective serotonin reuptake inhibitors. Furthermore, we parsed the overall predictive patterns into distinct components, revealing constitutive network constellations with generalizable SC-FC co-variation and treatment-specific traits. The TOMMF framework, with its demonstrated generalizability and high interpretability, offers enhanced predictive power for antidepressant responses with higher trustability, paving the way for precision medicine in MDD and providing a valuable tool for future multimodal neuroimaging biomarker studies.

Generalizability is crucial for translating biomarker discoveries into clinical practice. In this work, we evaluated our treatment response prediction models using an independent cohort of escitalopram-medicated patients. Our results suggest that the sertraline biomarker may generalize to predict escitalopram response, supporting the previous finding of a potential shared biomarker for serotonin reuptake inhibitors^29^. However, it is important to acknowledge that sertraline and escitalopram may still differ in their specific mechanisms of action. Our generalizability analysis shows that the sertraline response biomarker explains a smaller proportion of variance in escitalopram response, echoing with previous studies that neuroimaging biomarkers can differentiate responses to these drugs^9, 30^. This is consistent with sertraline’s additional effects on dopamine receptors^31^ and escitalopram’s binding to the allosteric site of serotonin transporters^32^. While drug-specific biomarkers may offer more accurate treatment response predictions and thus improving drug recommendations, a general biomarker could be valuable for identifying non-responders to first-line antidepressants. This would allow clinicians to expedite alternative treatment strategies without relying on trial-and-error, facilitating early intervention and personalized care.

Multimodal fusion is a technique emergingly employed by neuroimaging-based psychiatric studies, aiming to deepen our understanding of psychopathological neural alterations^33–36^ and improve predictability in psychiatric diagnosis^37–39^ and treatment effect^40^. Our innovative approach incorporates prediction target guidance into the multimodal fusion process, yielding substantial benefits over unsupervised fusion for downstream prediction tasks. As a recent study showed that the general neuroimaging variability across subjects mostly represent subclinical heterogeneity^41^, the target-oriented latent features ensured its relevance to antidepressant responses. Notably, the learning objective minimized the distance between canonical dimensions of SC and FC. Therefore, the latent features essentially reflect the information in agreement, thus identifying the association between SC and FC. Importantly, TOMMF’s linear basis simplifies the complexity, enhancing generalizability and facilitating neurophysiological interpretation of identified biomarkers. Furthermore, the L0-regularization achieved a high degree of orthogonality in latent features without explicit constraints, achieving data-driven dimensionality and subdimension identification in the overall predictive patterns, which elucidates the psychopharmacological intricacies of MDD.

Interestingly, our findings suggest that SC is more informative for predicting sertraline response, while FC is more indicative of placebo response. Conceptually, SC reflects the stable and enduring anatomical brain structure, while FC signifies the more adaptable active interactions between brain regions^17^. As previous studies have noted a significantly higher relapse rate with placebo compared to antidepressant medications after treatment discontinuation following remission^42–44^, we speculate that the greater variation in SC may contribute to the relatively persistent sertraline response, whereas the intermittent placebo response is associated with greater variation in FC. Due to the design of the clinical trial, we were unable to examine the heterogeneity in relapse using the EMBARC dataset. Additional research is warranted to provide further evidence for this theory, including investigating whether patients experiencing relapse earlier exhibit greater FC alteration compared to those experiencing relapse later or without relapse, irrespective of the treatment received. Its generalizability to other antidepressant medications should also be investigated.

Leveraging the innovative TOMMF framework, we identified neuroimaging biomarkers for antidepressant response on a multimodal basis. Notably, while sertraline and placebo responses exhibited distinct biomarkers, the precuneus emerged as a consistently significant predictor for both treatments. This underscores the crucial role of the precuneus in modulating antidepressant effects, aligning with previous findings indicating its abnormalities in MDD patients^45^ and predictive value for treatment outcome^11, 46^. Our study also provided insights into the contribution of subcortical regions to sertraline response, a facet not extensively explored in prior individual-level antidepressant response prediction studies^11, 46^. Specifically, we found functional connectivity involving striatum is particularly important for sertraline response, echoing with previous findings that FC involving striatum substantially associates with MDD symptoms^47^ and responses to repetitive transcranial magnetic stimulation^48^. Our models did not attribute as much importance to hippocampal structural abnormalities for antidepressant response, despite the hippocampus being consistently identified as a biomarker for MDD diagnosis^49^. This highlights the unique information provided by SC compared with anatomical volume of brain regions and suggests a possible discrepancy between the psychopathology of psychopharmacology of MDD. Moreover, the subcortical regions exhibited less substantial contribution in placebo response, in line with a recent finding highlighting differences in brain gradient between sertraline and placebo responses^46^. Importantly, our biomarker findings discerned between the contributions of SC and FC, providing novel insights into the respective roles of persistent and adaptable neural connections in treatment efficacy. Inspired by these findings, we advocate for future research into how persistent and adaptable neural connectivity influences antidepressant response and how therapeutics can facilitate the transition from transient treatment-induced neurophysiological changes to persistent ones.

Finally, the construction of latent features effectively disentangled the overarching predictive patterns for antidepressant response into distinct components. Our findings unveiled three distinct network constellations (i.e., clusters of networks) that synergistically informed the prediction of antidepressant response. These constellations, including the default-mode regulatory, affective, and sensory processing constellations, encompassed a spectrum from intrinsic to extrinsic functions and demonstrated a high degree of independence from each other. As validated by their robust SC-FC co-variation and generalizability across treatment arms, these network constellations offered novel insights into the intricate psychopharmacology of MDD. They also suggested personality traits and behavioral/cognitive task performance at baseline as potential indicators of treatment outcome. Specifically, the flanker inference on accuracy and choice reaction time at baseline linked to placebo response, whereas no examined task performance exhibited significant association with sertraline response, both aligning with a previous study^50^. To further validate and extend our discoveries, we encourage future studies to scrutinize the heterogeneity in response to other antidepressant therapeutics and explore potential distinctions between pharmacological and pathological aspects^49^.

In summary, we present a novel, adaptable, and highly explainable multimodal framework capable of predicting response to sertraline and placebo. The identified sertraline response biomarker is replicable on an independent escitalopram-medicated cohort, demonstrating its high trustability and suggesting an overlap of psychopharmacological mechanisms across selective serotonin reuptake inhibitors. Beyond the biomarker identification, our framework illuminates the intricate interplay between data modalities and discerns distinct components within the overall predictive patterns, which establishes it as a promising tool for investigating psychiatric disorders with multimodal neuroimaging data. Our key observation emphasizes that sertraline and placebo responses correspond to greater variation in SC and FC, underscoring the significance of persistent neurophysiological changes for optimal treatment outcomes. Furthermore, we have delineated three network constellations constituting the overarching antidepressant response biomarkers, which unveil potential distinct aspects within the complex psychopharmacology of MDD. These novel insights serve as inspiration for future research, advocating the development of therapeutics that induce transient changes to persistent alterations and address each constitutive neural circuitry within the comprehensive psychopharmacological pathways of MDD. With its demonstrated generalizability and high explainability, our findings hold promise for enabling neuroimaging-guided precision medicine and provide new insights for MDD that may foster the development of more efficacious antidepressant treatments.

## Methods

### Participants

#### Dataset 1 – The EMBARC Cohort

The <u>E</u>stablishing <u>M</u>oderators and <u>B</u>iosignatures of <u>A</u>ntidepressant <u>R</u>esponse in <u>C</u>linical Care (EMBARC) recruited 296 MDD patients and randomly assigned these subjects into the sertraline and placebo treatment arms^24^. After excluding disqualified/withdrawn subjects and the subjects with missing or low-quality neuroimaging data, we included 103 sertraline-medicated and 120 placebo-medicated MDD patients aged between 18 and 65 in our analyses (SFig. 4). The EMBARC study was conducted according to the FDA guidelines and the Declaration of Helsinki and approved by Institutional Review Board of each clinical site. All participants have given signed informed consent and agreement to all procedures prior to entry. The inclusion criteria of EMBARC study included: 1) MDD as the primary diagnosis by the Structured Clinical Interview for DSM-IV Axis I Disorders^51^, 2) Quick Inventory of Depressive Symptomatology score ≥ 14, 3) a MDD episode onset prior to age 30, either a chronic recurrent episode (duration ≥ 2 years) or recurrent MDD (≥ two lifetime episodes), and 4) no antidepressant failure during the current episode. The exclusion criteria of EMBARC study included: 1) ongoing pregnancy or breastfeeding, 2) being sexually active without contraception, 3) lifetime history of psychosis or bipolar disorder, 4) substance dependence in last 6 months or substance abuse in last 2 months, 5) unstable psychiatric or general medical conditions that require hospitalization, 6) study medication contraindication, 7) clinically significant laboratory abnormalities, 8) history of epilepsy or conditions requiring an anticonvulsant, 9) electroconvulsive therapy, vagal nerve stimulation, TMS or other somatic treatments in the current episode, 10) ongoing medication intake (including but not limited to antipsychotics and mood stabilizers), 11) ongoing psychotherapy, 12) significant suicide risk, and 13) unresponsiveness to any antidepressant at adequate dose and duration in the current episode.

For patients in either treatment arm, an eight-week course of sertraline or placebo was administered. Stratification was enforced by site, depression severity, and chronicity for the randomization of treatment arm. The dosing of medications started at 50mg and increased to a maximum of 200mg given patient’s tolerance and unresponsiveness to lower dosing. The treatment response was evaluated using HAMD_17_. Unavailable endpoint HAMD_17_ scores were imputed from subjects’ intermediate treatment response where possible to maximally retain the sample size. Additionally, neuroimaging data (including diffusion and functional MRI) were scanned for MDD patients before the treatment.

#### Dataset 2 – The CAN-BIND-1 Cohort

The initial samples of <u>Can</u>adian <u>B</u>iomarker Integration <u>N</u>etwork in <u>D</u>epression (CAN-BIND)-1 study included 196 MDD patients^52^. The inclusion criteria^53^ were: 1) outpatients aged 18 to 60, 2) experiencing a major depressive episode diagnosed by DSM-IV Text Revision and confirmed by the Mini International Neuropsychiatric Interview, 3) depressive episode duration ≥ 3 months, 4) Montgomery-Åsberg Depression Rating Scale (MADRS) score ≥ 24, and 5) sufficient English proficiency to complete interviews and self-report questionnaires. The exclusion criteria included: 1) bipolar I or II disorder, 2) primary diagnosis of psychiatric disorders other than MDD, 3) significant personality disorder that might interfere with study participation (e.g., borderline, antisocial), 4) high suicidal risk, 5) substance dependence or abuse in the past six months, 6) significant neurological disorders, head trauma, or other unstable medical conditions, 7) ongoing pregnancy or breastfeeding, 8) psychosis in the current episode, 9) high risk for hypomanic switch, 10) resistance to at least four antidepressant medications at therapeutic dosages, 11) previous unresponsiveness or contraindication to escitalopram or aripiprazole, 12) psychological treatment within past three months with intent to continue, and 13) contraindications to MRI. The CAN-BIND-1 study was approved by the ethics committees of each participating clinical site. All eligible participants have provided written informed consent for all study procedures at the screening visit.

For patients who had taken psychoactive medications, a wash-out period of at least five half-lives of medication was required. Subsequently, all MDD patients were treated with 10-20 mg/day escitalopram for eight weeks. Treatment response was measured as the change in MADRS score from pre- to post-treatment. We calculated the HAMD_17_ score estimated by equipercentile for a more direct comparison between results obtained with the EMBARC and CAN-BIND-1 cohorts (MADRS ≈ 1.3 × HAMD_17_)^54^. Baseline fMRI and DTI scans were also collected.

### MRI Acquisition

#### Functional MRI Acquisition

In the EMBARC study, resting-state fMRI data were collected using a single-shot gradient echo-planar pulse sequence with a duration around 5 minutes. While different scanners were used at four clinical sites (Columbia University: General Electric 3T; Massachusetts General Hospital: Siemens 3T; University of Texas Southwestern Medical Center: Philips 3T; University of Michigan: Philips 3T), acquisition parameters were standardized. Key parameters included: 2000 msec repetition time, 28 msec echo time, 90° flip angle, 64 × 64 matrix size, 3.2 × 3.2 x 3.1 mm^3^ voxel size, 39 axial slices, and 180 image volumes.

In the CAN-BIND-1 study, resting-state fMRI data were acquired over a 10-minute scan using a whole-brain T2*-sensitive blood-oxygen-level-dependent (BOLD) echo planar imaging sequence. Participants were instructed to remain still with their eyes open, focusing on a fixation cross^55^ throughout the scan. Key acquisition parameters, consistent across study sites, included a repetition time of 2000 msec, an echo time of 30 msec, and voxel dimensions of 4 mm × 4 mm × 4 mm^56^. Scanners varied by site (GE 3T for Toronto Western/Toronto General Hospital, Centre for Addiction and Mental Health, McMaster University, and University of Calgary; Philips 3T for University of British Columbia; Siemens 3T for Queen’s University). Notably, the University of British Columbia and Queen’s University used a significantly lower pixel bandwidth for fMRI scans^56^, potentially resulting in low signal-to-noise ratio of their data.

#### Diffusion MRI Acquisition

The same scanners used for fMRI acquisition were employed for diffusion MRI data collection in both cohorts. For the EMBARC cohort, diffusion MRI parameters varied by clinical site: Columbia University used a repetition time of 6 msec, echo time of 2.4 msec, 9° flip angle, and a 5-minute scan duration; Massachusetts General Hospital used 2.3 msec repetition time, 2.54 msec echo time, 9° flip angle, and 4.3-minute duration; University of Texas Southwestern Medical Center used 8 msec repetition time, 3.7 msec echo time, 12° flip angle, and 4.24-minute duration; University of Michigan used 8.1 msec repetition time, 3.7 msec echo time, 12° flip angle, and 5.29-minute duration.

In the CAN-BIND-1 study, DTI data were collected using a single-shot spin-echo echo planar imaging sequence^56^. Diffusion sensitizing gradients were applied in 31 non-collinear directions (b = 1000 s/mm²) and 6 volumes with b = 0 s/mm². The University of British Columbia and Queen’s University also used a significantly lower pixel bandwidth for DTI scans^56^, yielding more distorted images. We therefore excluded all subjects from these two study sites to improve the data quality for replication analysis.

### MRI Preprocessing

#### Diffusion MRI Preprocessing

Images with a b-value less than 100 s/mm^2^ were first designated as b=0 images. A denoising procedure was then implemented using the MRtrix3’s method^57^ with a 5-voxel window. Subsequently, correction for B1 field inhomogeneity was conducted using the dwibiascorrect function from MRtrix3 with the N4 algorithm incorporated^58^. Afterward, the mean intensity of the DWI series was adjusted to calibrate the mean intensity of the b=0 images across each individual DWI scanning sequence. FSL’s eddy was employed for head motion and Eddy correction^59^, with a q-space smoothing factor of 10, 5 iterations, and 1000 voxel-based hyperparameter estimation. Linear first- and second-level models were employed to characterize Eddy current-related spatial distortion and q-space coordinates were assigned to the respective shells. Field offset was separated from subject movement. Post-eddy, Eddy’s outlier replacement^60^ was conducted. Data were further organized by slices, ensuring the inclusion of values from slices with a minimum of 250 intracerebral voxels. Outlier groups that deviated by more than 4 standard deviations from the prediction were replaced with imputed values.

#### Functional MRI Preprocessing

The acquired resting-state fMRI data were preprocessed using the fMRIPrep pipeline^61^. Specifically, the T1-weighted images were corrected for intensity nonuniformity and stripped skull. Spatial normalization was conducted via nonlinear registration with the T1-weighted reference^62^. Brain tissue was segmented from the reference brain-extracted T1-weighted image^63^. The fieldmap information was used to correct distortion in low- and high-frequency components caused by field inhomogeneity. Subsequently, a corrected echo-planar imaging reference was obtained from a more accurate co-registration with the anatomical reference. The blood-oxygen-level-dependent (BOLD) reference was then transformed to the T1-weighted image with a boundary-based registration method^64^. Automatic removal of motion artifacts using independent component analysis^65^ was performed on the preprocessed BOLD time-series after removal of non-steady-state volumes and spatial smoothing with an isotropic Gaussian kernel of 6 mm full-width half-maximum. Lastly, quality control was enforced to remove data with high head motion (>0.5 mm motion framewise displacement or BOLD signal displacements > 0.5%)^66, 67^.

### Connectivity Feature Calculation

#### Structural Connectivity Calculation

We calculated the structural connectivity (SC) of all subjects using DSI studio program (https://dsi-studio.labsolver.org). Specifically, we first created a white matter mask with thresholding. Afterward, the spin distribution function of each voxel of masked image was estimated using generalized q-sampling imaging (GQI) reconstruction^68^ in the T1-weighted space with a diffusion sampling length ratio of 1.25. The tensor metrics were calculated using DWI with b-value lower than 1750 s/mm². The whole brain fibers were reconstructed using a modified streamline deterministic tracking algorithm^69^ with augmented tracking strategies^70^ and whole brain seeding strategy. The parameters were set as angular threshold 45°, step size 1 mm, fiber length extent 20–300 mm, and maximum subvoxel search seeds 1,000,000. In each tracking iteration, a random voxel coordinate within the whole brain seed was selected and fiber tracking algorithm started from the coordinate and tracks in two directions until the fiber reaches the brain border. Tracks with lengths shorter than 30 or longer than 200 mm were discarded. Finally, we extracted SC by calculating the counts of connected fibers linked pair-wise ROIs defined from the Schaefer atlas of 100 parcels^25^ and subcortical parcellations^26, 27^. The 35 subcortical ROIs include striatal and cerebellar regions, amygdala regions, anterior and posterior hippocampal regions, and the thalamus. We excluded the SC which do not exist in real brain or only exist in a proportion of subjects (< 67%) to improve the stability of predictive patterns across cross-validation folds. Lastly, the fiber numbers were log-transformed to enhance normality.

#### Functional Connectivity Calculation

Functional connectivity (FC) was extracted based on the same cortical and subcortical ROIs as used for structural connectivity extraction. The preprocessed fMRI data were averaged into 135 ROI-level time series. For each subject, FC was calculated as the Pearson’s correlation coefficient between the time series of each pair of ROIs. Fisher’s r-to-z transformation was implemented to ensure the normality of the FC features. For subjects with two fMRI scans, FC features were respectively calculated for each scan, and SC features were duplicated to form two multimodal samples. Samples corresponding to the same subject were never separated in the cross-validation procedure to prevent information leakage.

### The TOMMF framework (Multimodal-based Regression Model with L0-regularization)

To integrate information from multiple data modalities for improvement in prediction performance, we developed a framework that combines the multimodal fusion and regression task (Fig. 1a). Additionally, because the neural connectivity features outnumbered the sample size of subjects, sparsity constraint is introduced to control the model complexity thus alleviating the potential overfitting issue. Notably, here we utilized L0-regularization as the sparsity constraint for its non-shrinking variance and natural interpretability (see **Appendix B** for details), as compared with the commonly used L1-regularization (a.k.a. LASSO). We employed a technique named continuous sparsification^23^ to realize the optimization of the non-convex and non-differentiable L0-regularization (see **Appendix A** for details).

The overall loss function of the TOMMF framework can be formularized as:

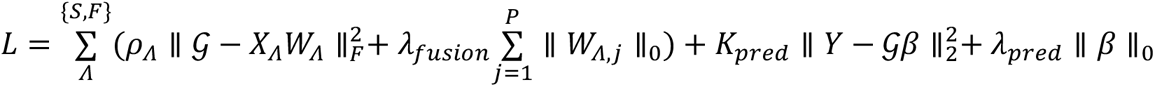

The notation *Λ* denotes the set of data modalities, in our case it contains the structural (S) and functional (F) modalities. The multimodal fusion term 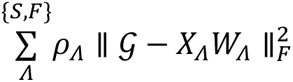 aims to construct a latent space 𝒢 that represents the information in agreement across data modalities, where the modality importance ratio *ρ_Λ_* is a hyperparameter that controls the relative contribution of each data modality. Incorporating *ρ_Λ_* into the multimodal fusion term enables the framework to lean the latent space toward the data modality that is more informative for a particular prediction target. Remarkably, as we minimize this multimodal fusion term, the distance between *X_S_W_S_* and *X_F_W_F_* is also minimized. Therefore, the multimodal fusion step also identifies canonical association between SC and FC features.

The prediction task term 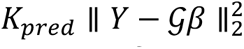 is a standard loss for prediction residue, where *K_pred_* modulates the relative importance of multimodal fusion and prediction loss. Both multimodal fusion and prediction task are regularized by the L0-norm of their feature weights (*W_S_*, *W_F_*, and *β*). This L0-regularization endows the framework with two significant characteristics – data-driven dimensionality and pseudo-exclusive compositions of the latent features (Fig. 1b, see **Appendix B** for details). These unique properties enable the framework to determine the number of features in the latent space based on their informativeness instead of an empirical percentage, as well as significantly enhance the interpretability of the latent features.

Additionally, by incorporating both multimodal fusion and prediction loss, our approach directs the fusion process with guidance from target – the optimal latent space 𝒢 is dependent on the target variable *Y*. This crucial relationship ensures that the latent space is tailored to the prediction target, resulting in distinct features for sertraline and placebo responses. In contrast to unsupervised multimodal fusion techniques (Fig. 2c, 3c), our proposed TOMMF framework conducts multimodal fusion in an unsupervised manner, enhancing the relevance of latent features and, thereby improving the prediction performance. With both multimodal fusion and prediction model relying on linear transformations, the overall predictive pattern from original SC and FC features to the antidepressant response is also linear. As a result, the TOMMF framework offers excellent end-to-end interpretability. Specifically, the equation for generating prediction *Ỹ* based on original SC and FC features (*X_S_* and *X_F_*) can be formularized as (see **Appendix A** for details):

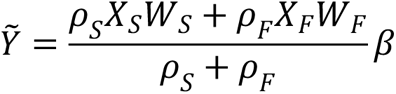

We can observe from the above formula that 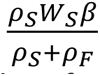 and 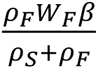 are the linear transformation matrices for the original SC and FC features, therefore granting the linear interpretability of the predictive biomarkers for antidepressant response.

### Generalization Analysis of Multimodal Treatment Response Biomarkers

To assess the generalizability of identified treatment response biomarkers, we directly applied the sertraline and placebo response prediction models on the CAN-BIND-1 cohort. Two study sites with significantly different MRI acquisition protocols that might lead to low data quality were excluded from the analysis (see **MRI Acquisition** for details). The final model for each treatment was calculated as the average of models in 100 cross-validation folds. No model re-training or fine-tuning was conducted. To leverage the longer fMRI scans to improve the data quality, fMRI data of the CAN-BIND-1 cohort were split into two sessions of equal length for FC calculation. An equipercentile-based conversion was employed to transform predicted HAMD17 change to symptom relief measured by MADRS^54^.

### Network Constellations

The network constellations were constructed to represent the distinct dimensions in the common space (Fig. 1c). To enhance their interpretability, we leveraged the dissection of functional networks which partition the brain into functional modules^71^. Specifically, we quantified the network components within biomarkers by summing ROI-level connectivity features involving a particular pair of networks, weighted by their coefficients from the antidepressant response prediction model. We calculated these components for SC and FC, respectively. Afterwards, we constructed network constellations by parsimoniously aggregating network components with the highest cross-modality correlations. Notably, SC and FC components for the same network were not considered equivalent. To derive psychopharmacological components with high generalizability and reliability, the network constellations were constructed based on sertraline response biomarker and then validated by the placebo response biomarker.

### NEO Personality Traits and Cognitive Tasks

We aimed to explore the relationship between personality traits and the network constellation components identified within antidepressant response biomarkers, building on recent findings of extensive associations between personality traits and psychopathology^28^. Personality traits were assessed using the NEO-five factor inventory, including agreeableness, neuroticism, extraversion, openness, and conscientiousness in MDD patients, as part of the EMBARC study^24^.

Additionally, behavioral/cognitive task performance has shown substantial associations with symptom profiles and treatment responsiveness in MDD^24^. The EMBARC study conducted five cognitive tasks, which we analyzed in this study. These tasks include the A-not-B task, choice reaction, the flanker test, the probabilistic reward task, and the word fluency test. Specifically, we examined patients’ accuracy in the A-not-B task, choice reaction time, Flanker inference on accuracy and reaction time, response bias in the probabilistic reward task, and the total number of valid words in the word fluency test. To identify potential pre-treatment biomarkers, we included only the cognitive task performance data collected at baseline.

### Statistical Tests

#### Comparing Intraclass Correlation Coefficients

To compare the intraclass correlation coefficients (ICCs) of predictive patterns derived from unimodal and multimodal frameworks, we performed two-sample z-test on the z-scores of ICCs. Specifically, Fisher z-transformation was first conducted on ICCs 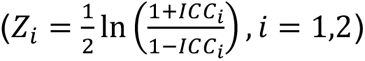 to ensure the normality of variables^72^. The z-statistic for two-sample z-test can be formularized as 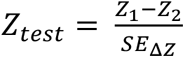, where 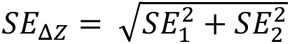 is the standard error of the difference between means and *SE_i_* (*i* = 1,2) is the standard error of each mean. Given the approximated standard error of Fisher’s z^73^: 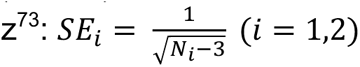, we can further formularize the z-statistic as:

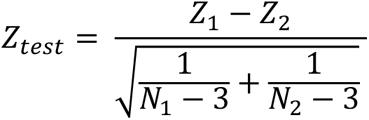

*N*_1_and *N*_2_ are the sample sizes for the two ICCs, which are the numbers of total connectivity features. Essentially, we examined the importance of each connectivity feature with the models in each of the cross-validation folds as judges. The p-value corresponding to the z-statistic can be found based on standard normal distribution.

## Data Availability

The EMBARC data are publicly available through the National Institute of Mental Health Data Archive (NDA) (https://nda.nih.gov/edit_collection.html?id=2199).

## Data Availability

The EMBARC cohort is publicly available through the National Institute of Mental Health Data Archive (NDA) (https://nda.nih.gov/edit_collection.html?id=2199). The CAN-BIND-1 cohort is available under a data use agreement with Brain-CODE, based at the Ontario Brain Institute (https://www.braincode.ca/content/canadian-biomarker-integration-network-depression-can-bind-0).

## Code Availability

All analyses were conducted in MATLAB (version R2022b) and the code will be made available upon the publication of this paper.

## Acknowledgements

This work was supported by NIH grant nos. R01MH129694, R21MH130956, R21AG080425, Alzheimer’s Association Grant (AARG-22-972541), and Lehigh University FIG (FIGAWD35), CORE, and Accelerator grants. Portions of this research were conducted on Lehigh University’s Research Computing infrastructure partially supported by NSF Award 2019035. G.A.F. was also supported by philanthropic funding and NIH grant nos. K23MH114023 and R01MH125886, and grants from the One Mind - Baszucki Brain Research Fund, the SEAL Future Foundation, and the Brain and Behavior Research Foundation. We would like to acknowledge the individuals and organizations that have made data available for this research, including CAN-BIND, the Ontario Brain Institute, the Brain-CODE platform, and the government of Ontario.

## Financial Disclosures

G.A.F. received monetary compensation for consulting work for SynapseBio AI and owns equity in Alto Neuroscience. A.E. reports salary and equity from Alto Neuroscience and equity in Akili Interactive and Mindstrong Health. C.J.K reports equity from Alto Neuroscience. The remaining authors declare no competing interests. C.N. is a consultant for ANeuroTech (division Anima BV), Janssen Research and Development, BioXcel Therapeutics, Engrail Therapeutics, Clexio Biosciences LTD, EmbarkNeuro, Galen Mental Health LLC, Goodcap Pharmaceuticals, ITI Inc, LUCY Scientific Discovery, Relmada Therapeutics, Sage Therapeutics, Senseye Inc, Precisement Health, Autobahn Therapeutics Inc, EMA Wellness, Skyland Trails, Denovo Biopharma, and the Brain & Behavior Research Foundation. C.N. owns the following patents: Method and devices for transdermal delivery of lithium (US 6,375,990B1), Method of assessing antidepressant drug therapy via transport inhibition of monoamine neurotransmitters by ex vivo assay (US 7,148,027B2), Compounds, Compositions, Methods of Synthesis, and Methods of Treatment (CRF Receptor Binding Ligand) (US 8,551, 996 B2). C.N. owns stock in Corcept Therapeutics Company, EMA Wellness, Precisement Health, Relmada Therapeutics, Signant Health, Galen Mental Health LLC, and Senseye Inc. The remaining authors have no conflicts of interest to declare.

## Notes

### Clinical Trial

NCT01407094

### Funding Statement

This work was supported by NIH grant nos. R01MH129694, R21MH130956, R21AG080425, Alzheimers Association Grant (AARG-22-972541), and Lehigh University FIG (FIGAWD35), CORE, and Accelerator grants. Portions of this research were conducted on Lehigh University Research Computing infrastructure partially supported by NSF Award 2019035. G.A.F. was also supported by philanthropic funding and NIH grant nos. K23MH114023 and R01MH125886, and grants from the One Mind - Baszucki Brain Research Fund, the SEAL Future Foundation, and the Brain and Behavior Research Foundation. We would like to acknowledge the individuals and organizations that have made data available for this research, including CAN-BIND, the Ontario Brain Institute, the Brain-CODE platform, and the government of Ontario.

### Author Declarations

The EMBARC data are publicly available through the National Institute of Mental Health Data Archive (NDA) (https://nda.nih.gov/edit_collection.html?id=2199). The CAN-BIND-1 cohort is available under a data use agreement with Brain-CODE, based at the Ontario Brain Institute (https://www.braincode.ca/content/canadian-biomarker-integration-network-depression-canbind-0)

### Summary of Updates

New validation results were added: Fig. 4 and section (Generalizability of Treatment Response Biomarkers on Independent Cohort).

## References

1. Lai, C.-H. Promising neuroimaging biomarkers in depression. Psychiatry investigation 16, 662 (2019).

2. Kambeitz, J. et al. Detecting neuroimaging biomarkers for depression: a meta-analysis of multivariate pattern recognition studies. Biological psychiatry 82, 330–338 (2017).

3. Buch, A.M. & Liston, C. Dissecting diagnostic heterogeneity in depression by integrating neuroimaging and genetics. Neuropsychopharmacology 46, 156–175 (2021).

4. Kraus, C., Kadriu, B., Lanzenberger, R., Zarate Jr, C.A. & Kasper, S. Prognosis and improved outcomes in major depression: a review. Translational psychiatry 9, 127 (2019).

5. Runia, N. et al. The neurobiology of treatment-resistant depression: a systematic review of neuroimaging studies. Neuroscience & Biobehavioral Reviews 132, 433–448 (2022).

6. Enneking, V., Leehr, E.J., Dannlowski, U. & Redlich, R. Brain structural effects of treatments for depression and biomarkers of response: a systematic review of neuroimaging studies. Psychological Medicine 50, 187–209 (2020).

7. Fu, C.H. et al. Neuroanatomical dimensions in medication-free individuals with major depressive disorder and treatment response to SSRI antidepressant medications or placebo. Nature Mental Health, 1–13 (2024).

8. Widge, A.S. et al. Electroencephalographic biomarkers for treatment response prediction in major depressive illness: a meta-analysis. American Journal of Psychiatry 176, 44–56 (2019).

9. Tozzi, L., Goldstein-Piekarski, A.N., Korgaonkar, M.S. & Williams, L.M. Connectivity of the cognitive control network during response inhibition as a predictive and response biomarker in major depression: evidence from a randomized clinical trial. Biological psychiatry 87, 462–472 (2020).

10. Wu, W. et al. An electroencephalographic signature predicts antidepressant response in major depression. Nature biotechnology 38, 439–447 (2020).

11. Zhao, K. et al. Individualized fMRI connectivity defines signatures of antidepressant and placebo responses in major depression. Molecular psychiatry, 1–10 (2023).

12. Geddes, J.R. et al. Relapse prevention with antidepressant drug treatment in depressive disorders: a systematic review. The Lancet 361, 653–661 (2003).

13. Kanai, T. et al. Time to recurrence after recovery from major depressive episodes and its predictors. Psychological Medicine 33, 839–845 (2003).

14. Breedvelt, J.J.F. et al. An individual participant data meta-analysis of psychological interventions for preventing depression relapse. Nature Mental Health (2024).

15. Honey, C.J. et al. Predicting human resting-state functional connectivity from structural connectivity. Proceedings of the National Academy of Sciences 106, 2035–2040 (2009).

16. Sui, J., Adali, T., Yu, Q., Chen, J. & Calhoun, V.D. A review of multivariate methods for multimodal fusion of brain imaging data. Journal of neuroscience methods 204, 68–81 (2012).

17. Zhang, Y.-D. et al. Advances in multimodal data fusion in neuroimaging: Overview, challenges, and novel orientation. Information Fusion 64, 149–187 (2020).

18. Tulay, E.E., Metin, B., Tarhan, N. & Arıkan, M.K. Multimodal neuroimaging: basic concepts and classification of neuropsychiatric diseases. Clinical EEG and neuroscience 50, 20–33 (2019).

19. Maglanoc, L.A. et al. Multimodal fusion of structural and functional brain imaging in depression using linked independent component analysis. Human brain mapping 41, 241–255 (2020).

20. Burke, M.J. et al. Placebo effects and neuromodulation for depression: a meta-analysis and evaluation of shared mechanisms. Molecular psychiatry 27, 1658–1666 (2022).

21. Pecina, M., Heffernan, J., Wilson, J., Zubieta, J.-K. & Dombrovski, A. Prefrontal expectancy and reinforcement-driven antidepressant placebo effects. Translational Psychiatry 8, 222 (2018).

22. Lii, T.R. et al. Randomized trial of ketamine masked by surgical anesthesia in patients with depression. Nature Mental Health, 1–11 (2023).

23. Savarese, P., Silva, H. & Maire, M. Winning the lottery with continuous sparsification. Advances in neural information processing systems 33, 11380–11390 (2020).

24. Trivedi, M.H. et al. Establishing moderators and biosignatures of antidepressant response in clinical care (EMBARC): Rationale and design. Journal of psychiatric research 78, 11–23 (2016).

25. Schaefer, A. et al. Local-Global Parcellation of the Human Cerebral Cortex from Intrinsic Functional Connectivity MRI. Cerebral Cortex 28, 3095–3114 (2017).

26. Buckner, R.L., Krienen, F.M., Castellanos, A., Diaz, J.C. & Yeo, B.T. The organization of the human cerebellum estimated by intrinsic functional connectivity. Journal of neurophysiology 106, 2322–2345 (2011).

27. Choi, E.Y., Yeo, B.T. & Buckner, R.L. The organization of the human striatum estimated by intrinsic functional connectivity. Journal of neurophysiology 108, 2242–2263 (2012).

28. Zhang, Y.-R. et al. Personality traits and brain health: a large prospective cohort study. Nature Mental Health, 1–14 (2023).

29. Korgaonkar, M.S., Goldstein-Piekarski, A.N., Fornito, A. & Williams, L.M. Intrinsic connectomes are a predictive biomarker of remission in major depressive disorder. Molecular psychiatry 25, 1537–1549 (2020).

30. Williams, L.M., Debattista, C., Duchemin, A., Schatzberg, A.F. & Nemeroff, C.B. Childhood trauma predicts antidepressant response in adults with major depression: data from the randomized international study to predict optimized treatment for depression. Translational psychiatry 6, e799–e799 (2016).

31. Damsa, C. et al. " Dopamine-dependent" side effects of selective serotonin reuptake inhibitors: a clinical review. Journal of Clinical Psychiatry 65, 1064–1068 (2004).

32. Xue, W. et al. Molecular mechanism for the allosteric inhibition of the human serotonin transporter by antidepressant escitalopram. ACS chemical neuroscience 13, 340–351 (2022).

33. Sui, J. et al. In search of multimodal neuroimaging biomarkers of cognitive deficits in schizophrenia. Biological psychiatry 78, 794–804 (2015).

34. Foti, D., Carlson, J.M., Sauder, C.L. & Proudfit, G.H. Reward dysfunction in major depression: Multimodal neuroimaging evidence for refining the melancholic phenotype. NeuroImage 101, 50–58 (2014).

35. Scheepens, D.S. et al. The link between structural and functional brain abnormalities in depression: A systematic review of multimodal neuroimaging studies. Frontiers in Psychiatry 11, 485 (2020).

36. Johnston, J.A. et al. Multimodal neuroimaging of frontolimbic structure and function associated with suicide attempts in adolescents and young adults with bipolar disorder. American journal of psychiatry 174, 667–675 (2017).

37. Shi, J., Zheng, X., Li, Y., Zhang, Q. & Ying, S. Multimodal neuroimaging feature learning with multimodal stacked deep polynomial networks for diagnosis of Alzheimer’s disease. IEEE journal of biomedical and health informatics 22, 173–183 (2017).

38. Liu, S. et al. Multimodal neuroimaging feature learning for multiclass diagnosis of Alzheimer’s disease. IEEE transactions on biomedical engineering 62, 1132–1140 (2014).

39. Lei, D. et al. Integrating machining learning and multimodal neuroimaging to detect schizophrenia at the level of the individual. Human brain mapping 41, 1119–1135 (2020).

40. Weigand, A. et al. Predicting antidepressant effects of ketamine: the role of the pregenual anterior cingulate cortex as a multimodal neuroimaging biomarker. International Journal of Neuropsychopharmacology 25, 1003–1013 (2022).

41. Tong, X. et al. Symptom dimensions of resting-state electroencephalographic functional connectivity in autism. Nature Mental Health (2024).

42. Kato, M. et al. Discontinuation of antidepressants after remission with antidepressant medication in major depressive disorder: a systematic review and meta-analysis. Molecular psychiatry 26, 118–133 (2021).

43. Hansen, R. et al. Meta-analysis of major depressive disorder relapse and recurrence with second-generation antidepressants. Psychiatric services 59, 1121–1130 (2008).

44. Williams, N., Simpson, A.N., Simpson, K. & Nahas, Z. Relapse rates with long-term antidepressant drug therapy: a meta-analysis. Human Psychopharmacology: Clinical and Experimental 24, 401–408 (2009).

45. Cheng, W. et al. Functional connectivity of the precuneus in unmedicated patients with depression. Biological Psychiatry: Cognitive Neuroscience and Neuroimaging 3, 1040–1049 (2018).

46. Tong, X. et al. Individual deviations from normative electroencephalographic connectivity predict antidepressant response. Journal of Affective Disorders (2024).

47. Gabbay, V. et al. Striatum-based circuitry of adolescent depression and anhedonia. Journal of the American Academy of Child & Adolescent Psychiatry 52, 628–641.e613 (2013).

48. Avissar, M. et al. Functional connectivity of the left DLPFC to striatum predicts treatment response of depression to TMS. Brain stimulation 10, 919–925 (2017).

49. Otte, C., et al. Major depressive disorder. Nature reviews Disease primers 2, 1–20 (2016).

50. Ang, Y.-S. et al. Exploration of baseline and early changes in neurocognitive characteristics as predictors of treatment response to bupropion, sertraline, and placebo in the EMBARC clinical trial. Psychological medicine 52, 2441–2449 (2022).

51. First, M.B., Spitzer, R.L., Gibbon, M. & Williams, J.B. (SCID-I/P New York, NY, USA:, 2002).

52. Nogovitsyn, N. et al. Hippocampal tail volume as a predictive biomarker of antidepressant treatment outcomes in patients with major depressive disorder: a CAN-BIND report. Neuropsychopharmacology 45, 283–291 (2020).

53. Lam, R.W. et al. Discovering biomarkers for antidepressant response: protocol from the Canadian biomarker integration network in depression (CAN-BIND) and clinical characteristics of the first patient cohort. BMC psychiatry 16, 1–13 (2016).

54. Leucht, S., Fennema, H., Engel, R.R., Kaspers-Janssen, M. & Szegedi, A. Translating the HAM-D into the MADRS and vice versa with equipercentile linking. Journal of affective disorders 226, 326–331 (2018).

55. Raichle, M.E. & Snyder, A.Z. A default mode of brain function: a brief history of an evolving idea. Neuroimage 37, 1083–1090 (2007).

56. MacQueen, G.M. et al. The Canadian biomarker integration network in depression (CAN-BIND): magnetic resonance imaging protocols. Journal of Psychiatry & Neuroscience: JPN 44, 223 (2019).

57. Veraart, J. et al. Denoising of diffusion MRI using random matrix theory. Neuroimage 142, 394–406 (2016).

58. Tustison, N.J. et al. N4ITK: improved N3 bias correction. IEEE transactions on medical imaging 29, 1310–1320 (2010).

59. Andersson, J.L.R. & Sotiropoulos, S.N. An integrated approach to correction for off-resonance effects and subject movement in diffusion MR imaging. Neuroimage 125, 1063–1078 (2016).

60. Andersson, J.L.R., Graham, M.S., Zsoldos, E. & Sotiropoulos, S.N. Incorporating outlier detection and replacement into a non-parametric framework for movement and distortion correction of diffusion MR images. Neuroimage 141, 556–572 (2016).

61. Esteban, O. et al. fMRIPrep: a robust preprocessing pipeline for functional MRI. Nature methods 16, 111–116 (2019).

62. Avants, B.B., Epstein, C.L., Grossman, M. & Gee, J.C. Symmetric diffeomorphic image registration with cross-correlation: evaluating automated labeling of elderly and neurodegenerative brain. Medical image analysis 12, 26–41 (2008).

63. Zhang, Y., Brady, J.M. & Smith, S. in Medical Imaging 2000: Image Processing, Vol. 3979 1126–1137 (SPIE, 2000).

64. Greve, D.N. & Fischl, B. Accurate and robust brain image alignment using boundary-based registration. Neuroimage 48, 63–72 (2009).

65. Pruim, R.H. et al. ICA-AROMA: A robust ICA-based strategy for removing motion artifacts from fMRI data. Neuroimage 112, 267–277 (2015).

66. Power, J.D., Barnes, K.A., Snyder, A.Z., Schlaggar, B.L. & Petersen, S.E. Spurious but systematic correlations in functional connectivity MRI networks arise from subject motion. Neuroimage 59, 2142–2154 (2012).

67. Power, J.D., Barnes, K.A., Snyder, A.Z., Schlaggar, B.L. & Petersen, S.E. Steps toward optimizing motion artifact removal in functional connectivity MRI; a reply to Carp. Neuroimage 76 (2013).

68. Yeh, F.C., Wedeen, V.J. & Tseng, W.Y. Generalized q-sampling imaging. IEEE Trans Med Imaging 29, 1626–1635 (2010).

69. Yeh, F.C., Verstynen, T.D., Wang, Y., Fernandez-Miranda, J.C. & Tseng, W.Y. Deterministic diffusion fiber tracking improved by quantitative anisotropy. PLoS One 8, e80713 (2013).

70. Yeh, F.C. Shape analysis of the human association pathways. Neuroimage 223, 117329 (2020).

71. Thomas Yeo, B., et al. The organization of the human cerebral cortex estimated by intrinsic functional connectivity. Journal of neurophysiology 106, 1125–1165 (2011).

72. Weinberg, R. & Patel, Y.C. Simulated intraclass correlation coefficients and their z transforms. Journal of Statistical Computation and Simulation 13, 13–26 (1981).

73. Fouladi, R.T. & Steiger, J.H. The Fisher transform of the Pearson product moment correlation coefficient and its square: Cumulants, moments, and applications. Communications in Statistics—Simulation and Computation® 37, 928–944 (2008).

