## Supplementary Material for "Optimizing Antidepressant Efficacy: Generalizable Multimodal Neuroimaging Biomarkers for Prediction of Treatment Response"

### Supplementary Information

**SFig. 1** Permutation tests for TOMMF-based antidepressant response prediction
**SFig. 2** Relative importance of SC and FC for antidepressant response prediction
**SFig. 3** Treatment response predictive patterns derived with single data modality
**SFig. 4** CONSORT flow diagram of the EMBARC clinical trial

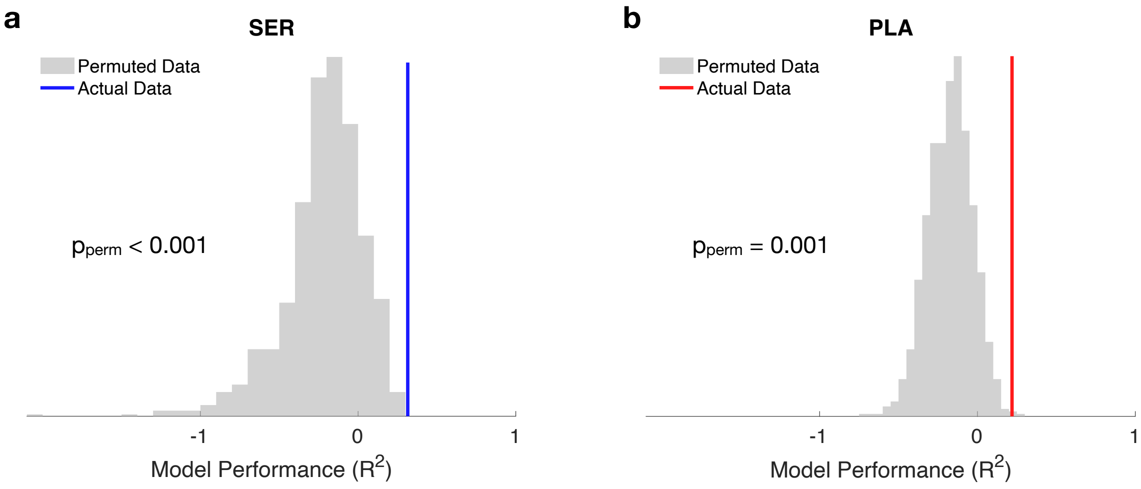


**SFig. 1** Permutation tests for TOMMF-based antidepressant response prediction. **a** Sertraline response prediction. **b** Placebo response prediction. Permutation tests are conducted by randomly permuting the prediction target of patients and are repeated 1000 times.

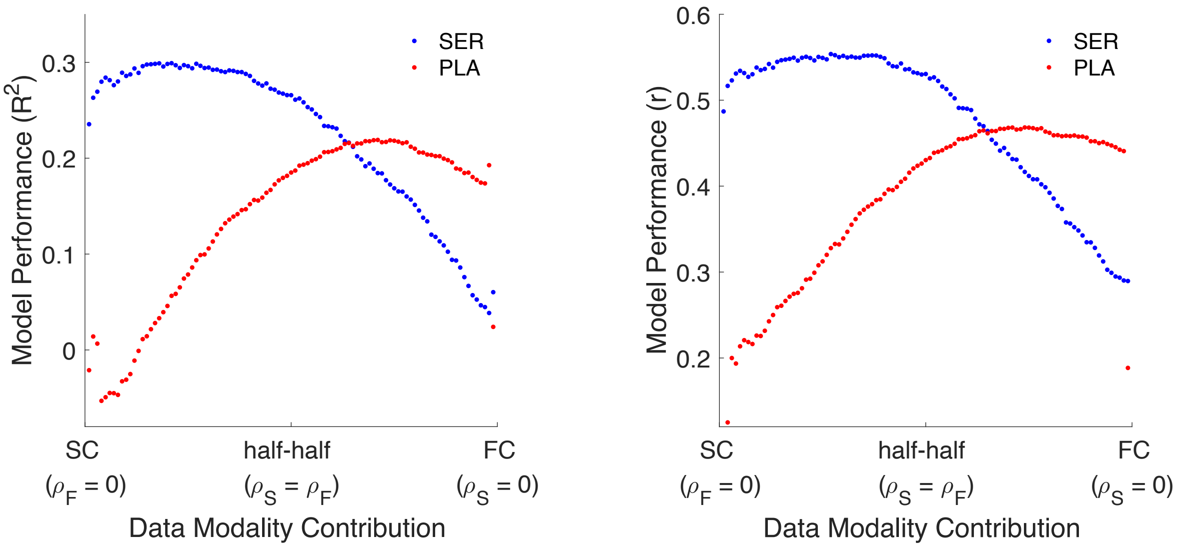


**SFig. 2** Relative importance of SC and FC for antidepressant response prediction. The SC features show stronger contribution to the sertraline response biomarker, while the FC features exhibit a more substantial influence on the placebo response biomarker. The modality importance parameters ($\rho_S$ and $\rho_F$) are tuned to evaluate the model performance for each importance ratio. The model performance is evaluated by both R-squared and the correlation coefficient (r) between actual and predicted values of antidepressant response.

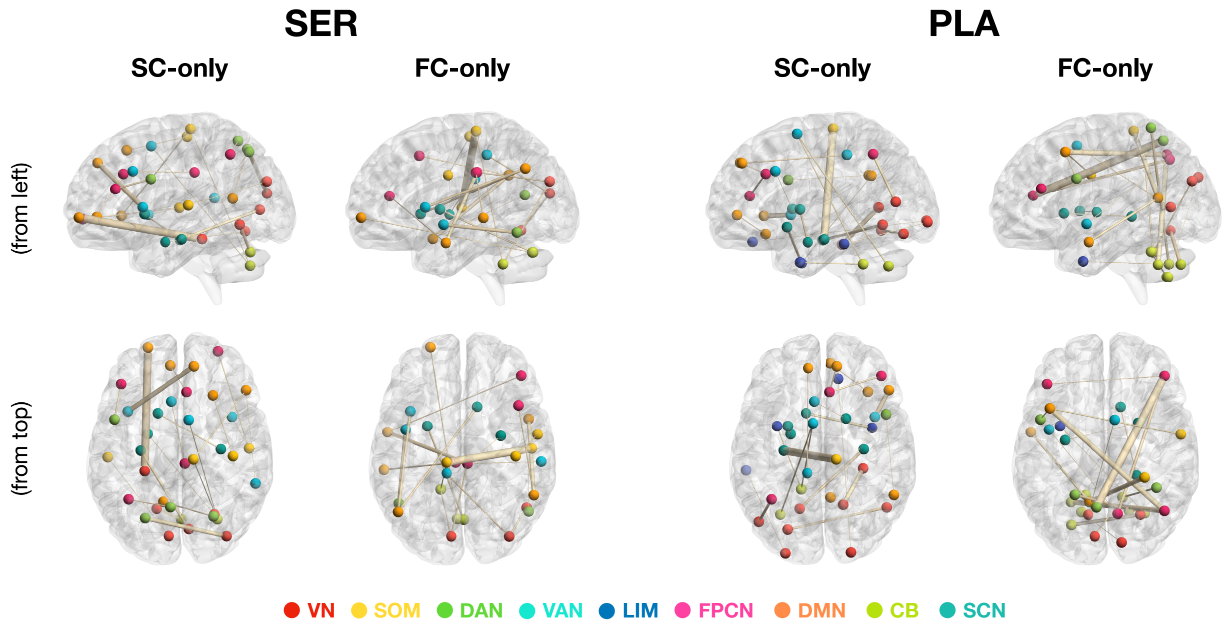


**SFig. 3** Treatment response predictive patterns derived with single data modality. LASSO regression is conducted with only SC or FC features for predicting sertraline (SER) and placebo (PLA) response. Overall, the unimodal-based predictive patterns exhibit significant alignment with multimodal-based predictive patterns, demonstrating the reliability of predictive biomarkers for antidepressant response.

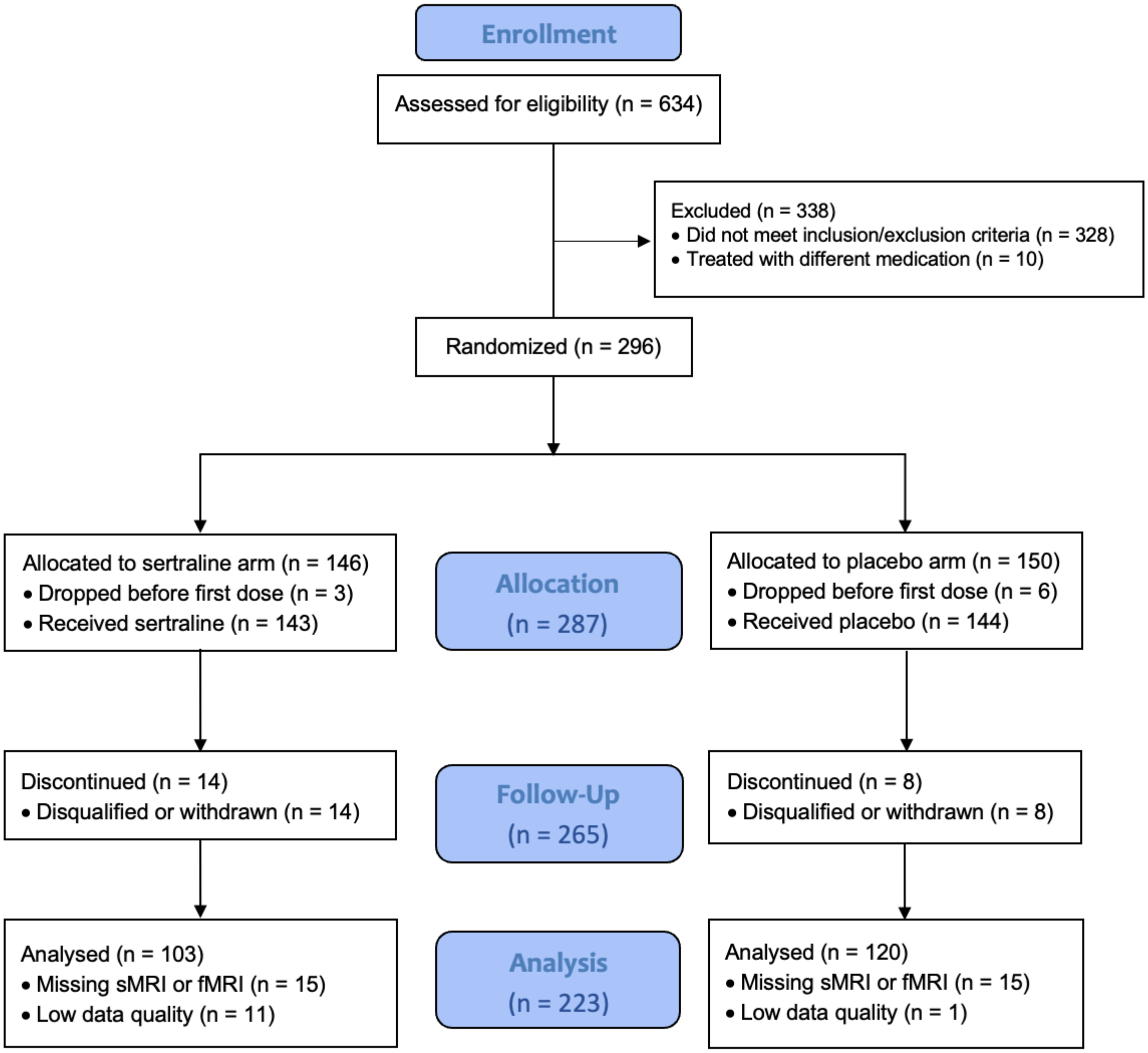


**SFig. 4** CONSORT flow diagram of the EMBARC clinical trial. The flow diagram shows the number of MDD patients who were randomized to treatment, received treatment, and had valid structural and functional MRI data available for the analyses. Patients who received partial treatment (less than 8 week) are included in the analyses with imputed treatment response.
