## Extended Data for "Optimizing Antidepressant Efficacy: Generalizable Multimodal Neuroimaging Biomarkers for Prediction of Treatment Response"

**Extended Data Fig. 1** Data driven dimensionality and dissimilarity of dimension compositions of the latent space.

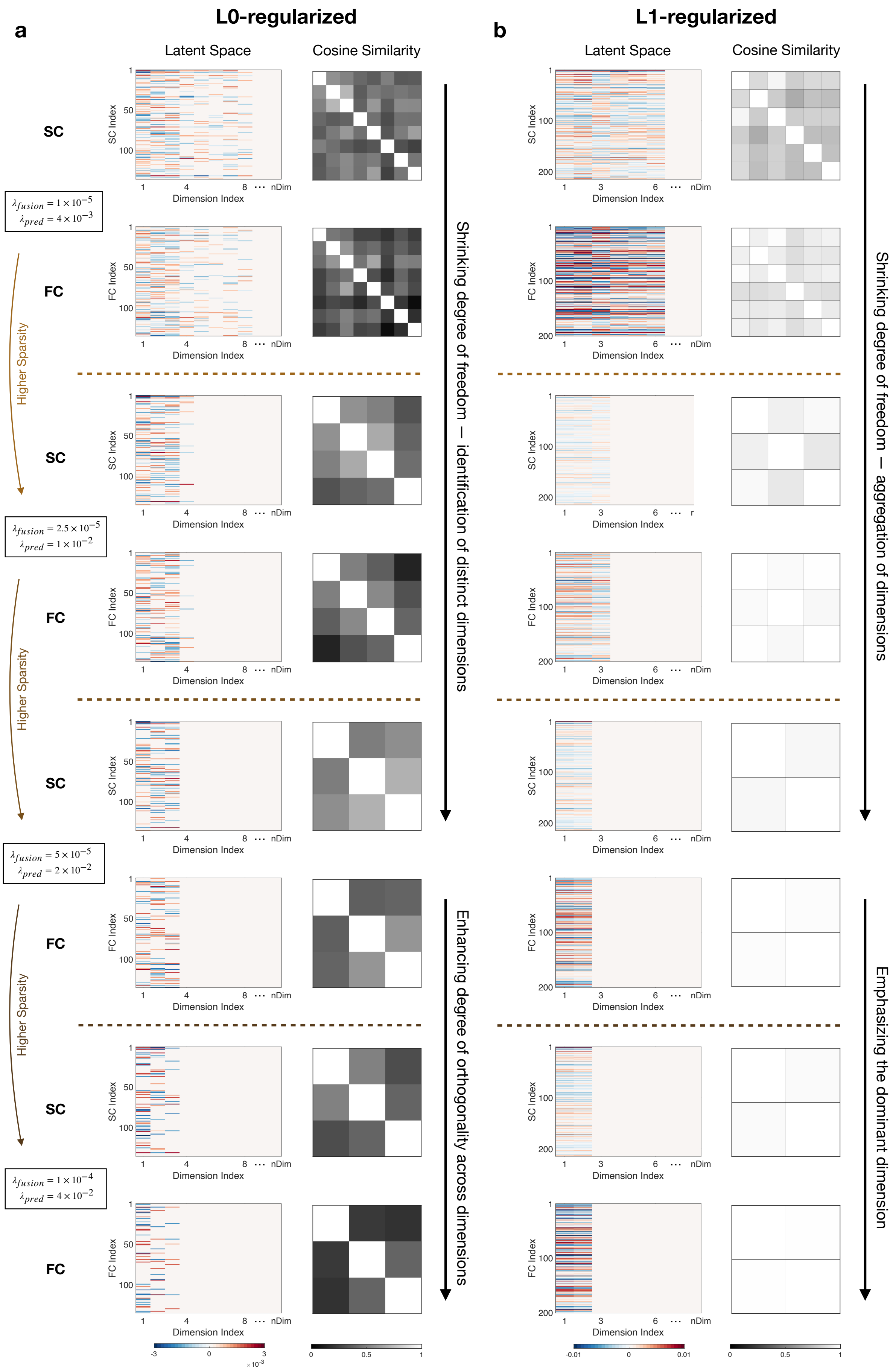

a
L0-regularized
b
L1-regularized
Latent Space
Cosine Similarity
SC
FC
SC Index
FC Index
Dimension Index
nDim
$\lambda_{fusion} = 1 \times 10^{-5}$
$\lambda_{pred} = 4 \times 10^{-3}$
$\lambda_{fusion} = 2.5 \times 10^{-5}$
$\lambda_{pred} = 1 \times 10^{-2}$
$\lambda_{fusion} = 5 \times 10^{-5}$
$\lambda_{pred} = 2 \times 10^{-2}$
$\lambda_{fusion} = 1 \times 10^{-4}$
$\lambda_{pred} = 4 \times 10^{-2}$
Higher Sparsity
Shrinking degree of freedom — identification of distinct dimensions
Enhancing degree of orthogonality across dimensions
Shrinking degree of freedom — aggregation of dimensions
Emphasizing the dominant dimension
$\times 10^{-3}$

**Extended Data Fig. 1** Data driven dimensionality and dissimilarity of dimension compositions of the latent space. As the degree of sparsity increases, the latent space dimensions derived from both the L0-regularized (**a**) and L1-regularized (**b**) framework first exhibit a shrinkage of degree of freedom (decrease in number of dimensions), demonstrating the data driven dimensionality property that the framework can judge on its own whether a feature is adequately informative to be included in the sparse latent space. However, while the latent dimensions derived with L0-regularization display decent dissimilarity with each other (as quantified by their pairwise cosine similarity), the dimensions derived with L1-regularization gradually converge. After the dimensionality reaches a relative stable number, the L0-regularized framework further enhance the degree of orthogonality across dimensions, while the L1-regularized counterpart further aggregates information from dimensions and emphasizes the dominant dimension. Together, the differences in the properties of latent space demonstrate the unique advantage of L0-regularized framework over its L1-regularized counterpart — the L0-regularization imposes penalty on the occurrence of features in dimensions, especially the features with contribution to multiple dimensions, therefore yielding distinct dimensions with decent dissimilarity with each other. The sparsity parameters ($\lambda_{fusion}$ and $\lambda_{pred}$) are harmoniously adjusted and kept same for L0- and L1-regularization. The colormap of latent space shows the weights of informative connectivity features with respect to each of the latent dimensions. The blankness in the rightmost part of latent space is the eliminated dimensions that are automatically set to zero vectors by the framework due to their inadequate informativeness. The latent space of one cross-validation fold is shown for each regularization paradigm as the representative example.
