## Appendix A for "Optimizing Antidepressant Efficacy: Generalizable Multimodal Neuroimaging Biomarkers for Prediction of Treatment Response"

### Appendix A — Optimization of the TOMMF framework

To derive the procedure to optimize the model for TOMMF, let us first revisit the overall loss function:

$$L = \sum_{\Lambda}^{\{S,F\}} \left(\rho_\Lambda \parallel \mathcal{G} - X_\Lambda W_\Lambda \parallel_F^2 + \lambda_{fusion} \sum_{j=1}^{P} \parallel W_{\Lambda,j} \parallel_0\right) + K_{pred} \parallel Y - \mathcal{G}\beta \parallel_2^2 + \lambda_{pred} \parallel \beta \parallel_0$$

The most common workaround to optimize L0-regularization is hard-thresholding, where in each iteration, the K % feature weights with smallest absolute values are set to 0 (K is a fixed number reflecting the desired sparsity level and/or prior knowledge about the proportion of informative features). However, hard-thresholding essentially imposes an extra constraint on the solution of loss function instead of solving the L0-regularized loss itself. Moreover, hard-thresholding requires a predetermined sparsity level, which prevents the adjustment of sparsity during the training process. Although this is typically not problematic in architectures with multilayer neural networks, where adjacent layers can compensate for the inflexibility of the sparsified layer, our framework requires an approach to optimize the sparsity level in a data-driven way for higher reliability and accuracy. Fortunately, we found the continuous sparsification technique[1] born for this purpose.

#### *Continuous Sparsification — Remodeling L0-regularization as Dichotomized Masks*

L0-regularization essentially imposes penalty on the inclusion of features. The inclusion/exclusion of features can be further modeled as masks applying on each of the feature weights — non-zero weights are with a mask of 1, whereas zero weights are with a mask of 0. With this concept, the equivalent feature weights for multimodal fusion and prediction task can be written as $W_\Lambda \odot M_\Lambda^*$ and $\beta \odot M_\beta^*$, where $M^*$ represents the masks and $\odot$ is element-wise multiplication. Thus, we can get $\parallel W_\Lambda \odot M_\Lambda^* \parallel_0 = \parallel M_\Lambda^* \parallel_1$ and $\parallel \beta \odot M_\beta^* \parallel_0 = \parallel M_\beta^* \parallel_1$.

Analogous to logistic regression, the continuous sparsification technique[1] approximates the dichotomized masks using a sigmoid function: $M^* \approx \sigma(\alpha M)$, where $\alpha$ is the parameter controlling the steepness of the sigmoid function $\sigma(\alpha M_\Lambda) = \frac{1}{1+e^{-\alpha M_\Lambda}}$. Given the fact that: $\lim_{\alpha \to \infty} \sigma(\alpha M) = H(M)$, where $H(\cdot)$ is the Heaviside step function, we can write $M^* = \lim_{\alpha \to \infty} \sigma(\alpha M)$.

Taken together, we can rewrite the loss function using the mask terms as:

$$L = \sum_{\Lambda}^{\{S,F\}} \left(\rho_\Lambda \parallel \mathcal{G} - X_\Lambda(\sigma(\alpha_W M_\Lambda) \odot W_\Lambda) \parallel_F^2 + \lambda_{fusion} \sum_{j=1}^{P} \parallel \sigma(\alpha_W M_{\Lambda,j}) \parallel_1\right) + K_{pred} \parallel Y - \mathcal{G}(\sigma(\alpha_\beta M_\beta) \odot \beta) \parallel_2^2 + \lambda_{pred} \parallel \sigma(\alpha_\beta M_\beta) \parallel_1$$

Notably, the regularization is not applied on the feature weights $W_\Lambda$ and $\beta$ anymore. Instead, the original L0-regularization is converted to L1-regularization on the masks, which is ready to be optimized. Furthermore, it is easy to see that the sigmoid function is always positive, making its L1-norm differentiable everywhere.

#### *Gradient Descent*

Overall, the loss function with mask terms can be optimized by gradient descent. We further found that the optimal value of $\mathcal{G}$ given other variables can be evaluated efficiently, therefore $\mathcal{G}$ was updated using its current optimal values instead of gradient descent. In fact, denoting $\gamma = \sigma(\alpha_\beta M_\beta) \odot \beta$, we can get:

$$\frac{\partial L}{\partial \mathcal{G}_{ij}} = 2(\rho_S + \rho_F)\mathcal{G}_{ij} + 2K_{pred}\gamma_j \mathcal{G}_i \gamma - (2\rho_S X_S W_S + 2\rho_F X_F W_F + 2K_{pred} Y \gamma^T)_{ij}$$

Given the value of other variables, the optimal value of $\mathcal{G}$ is reach when $\frac{\partial L}{\partial \mathcal{G}_{ij}} = 0$ for all i (rows) and j (columns). It is clear to see that each row of $\mathcal{G}$ constitutes a set of linear equations, yielding the optimal value of $\mathcal{G}$ as:

$$\mathcal{G} = (\rho_S X_S W_S + \rho_F X_F W_F + K_{pred} Y \gamma^T)(K_{pred}\gamma\gamma^T + (\rho_S + \rho_F)I)^{-1}$$

It is obvious that $K_{pred}\beta\beta^T + (\rho_S + \rho_F)I_P$ is Hermitian — it has an eigenvalue of $K_{pred}\gamma^T\gamma + \rho_S + \rho_F$ and a repeated eigenvalue of $\rho_S + \rho_F$ (repeated P-1 times), so it is always invertible. Evaluating the matrix inverse is generally not computationally efficient, but as $\gamma$ is a vector, this matrix inverse can be expressed in an explicit way:

$$(K_{pred}\gamma\gamma^T + (\rho_S + \rho_F)I)^{-1} = \frac{I}{\rho_S + \rho_F} - \frac{K_{pred}\gamma\gamma^T}{(\rho_S + \rho_F)^2 + (\rho_S + \rho_F)K_{pred}\gamma^T\gamma}$$

Hence, instead of evaluating its gradient descent, we directly set $\mathcal{G}$ to its current optimal value to accelerate the optimization procedure.

Together with the gradients of other variables, the basic optimization procedure can be formularized as ($\mu_W$, $\mu_M$, and $\mu_\beta$ are learning rates):

$$W_\Lambda^{t+1} = W_\Lambda^t - 2\mu_W \rho_\Lambda X_\Lambda^T (X_\Lambda(\sigma(\alpha_W M_\Lambda^t) \odot W_\Lambda^t) - \mathcal{G}^t) \odot \sigma(\alpha_W M_\Lambda^t)$$

$$M_\Lambda^{t+1} = M_\Lambda^t - \mu_M(2\rho_\Lambda X_\Lambda^T (X_\Lambda(\sigma(\alpha_W M_\Lambda^t) \odot W_\Lambda^t) - \mathcal{G}^t) \odot W_\Lambda^t + \lambda_{fusion}) \odot \sigma'(\alpha_W M_\Lambda^t)$$

$$\beta^{t+1} = \beta^t - 2\mu_\beta K_{pred}(\mathcal{G}^t)^T(\mathcal{G}^t(\sigma(\alpha_\beta M_\beta^{t+1}) \odot \beta^t) - Y) \odot \sigma(\alpha_\beta M_\beta^t)$$

$$M_\beta^{t+1} = M_\beta^t - \mu_M(2K_{pred}(\mathcal{G}^t)^T(\mathcal{G}^t(\sigma(\alpha_\beta M_\beta^{t+1}) \odot \beta^t) - Y) \odot \beta^t + \lambda_{pred}) \odot \sigma'(\alpha_\beta M_\beta^t)$$

$$\gamma^{t+1} = \sigma(\alpha_\beta M_\beta^{t+1}) \odot \beta^{t+1}$$

$$\mathcal{G}^{t+1} = (\rho_S X_S(\sigma(\alpha_\beta M_S^{t+1}) \odot W_S^{t+1}) + \rho_F X_F(\sigma(\alpha_\beta M_F^{t+1}) \odot W_F^{t+1}) + K_{pred} Y (\gamma^{t+1})^T)(\frac{I}{\rho_S + \rho_F} - \frac{K_{pred}\gamma^{t+1}(\gamma^{t+1})^T}{(\rho_S + \rho_F)^2 + (\rho_S + \rho_F)K_{pred}(\gamma^{t+1})^T\gamma^{t+1}})$$

Additionally, after T iterations, $\alpha_W$ and $\alpha_\beta$ are increased. Their increase follows exponential schedules, starting from 1. Remarkably, although the penalty on mask approximates L0-norm when $\alpha_W$ and $\alpha_\beta$ are large, during early stage of the training process, the regularization term is continuous and reflects L1-norm. This imposes constraints on the magnitude of the latent space $\mathcal{G}$ — penalty on W prevents the magnitude of $\mathcal{G}$ from being unboundedly high, penalty on $\beta$

prevents the magnitude of $\mathcal{G}$ from being unboundedly low, leading to balanced magnitude of optimized $\mathcal{G}$.

Lastly, it is worth noting that although we formularized $\rho_S$ and $\rho_F$ and two variables, they essentially modulate the ratio of relative importance of each data modality. Therefore, their degree of freedom is 1. Here, we kept $\rho_S + \rho_F$ as a constant and only searched for the best ratio between $\rho_S$ and $\rho_F$.

#### *Fine-tuning Variables with Fully Dichotomized Masks*

The last step of optimizing L0-regularized loss function is thresholding the pseudo-dichotomized masks to strictly dichotomized masks, and then running last iterations to fine-tune the model variables with inclusive masks (masks with value of 1). Essentially, this means replacing $\sigma(\alpha M)$ with $H(M)$, and then optimizing the non-regularized loss function with $M_\Lambda$ and $M_\beta$ being treated as constants:

$$L = \sum_{\Lambda}^{\{S,F\}} \rho_\Lambda \parallel \mathcal{G} - X_\Lambda(H(M_\Lambda) \odot W_\Lambda) \parallel_F^2 + K_{pred} \parallel Y - \mathcal{G}(H(M_\beta) \odot \beta) \parallel_2^2$$

With gradient descents, the optimization procedure can be formularized as:

$$W_\Lambda^{t+1} = W_\Lambda^t - 2\mu_W \rho_\Lambda X_\Lambda^T (X_\Lambda(H(M_\Lambda) \odot W_\Lambda^t) - \mathcal{G}^t) \odot H(M_\Lambda)$$

$$\beta^{t+1} = \beta^t - 2\mu_\beta K_{pred} (\mathcal{G}^t)^T (\mathcal{G}^t(H(M_\beta) \odot \beta^t) - Y) \odot H(M_\beta)$$

$$\gamma^{t+1} = H(M_\beta) \odot \beta^{t+1}$$

$$\mathcal{G}^{t+1} = (\rho_S X_S(H(M_S) \odot W_S^{t+1}) + \rho_F X_F(H(M_F) \odot W_F^{t+1}) + K_{pred} Y(\gamma^{t+1})^T)(\frac{I}{\rho_S + \rho_F} - \frac{K_{pred}\gamma^{t+1}(\gamma^{t+1})^T}{(\rho_S + \rho_F)^2 + (\rho_S + \rho_F)K_{pred}(\gamma^{t+1})^T \gamma^{t+1}})$$

Essentially, only the features with non-zero mask are learnable in this stage. As the selection of these features has already represented the implementation of L0-regularization, this final fine-tuning procedure does not include any regularization constraint. The weights of features with zero mask will not be adjusted, and those features will not contribute to the final prediction model.

#### *Evaluate Target on New Data*

Despite we solved $\mathcal{G}$ during the optimization procedure, it is not an available measure for new data. Moreover, the optimal $\mathcal{G}$ ia a function of both $W_\Lambda$ and $\beta$ at convergence, so a simple estimate of $\frac{\rho_S X_S W_S + \rho_F X_F W_F}{\rho_S + \rho_F}$ may not be accurate. Actually, $\frac{\rho_S X_S W_S + \rho_F X_F W_F}{\rho_S + \rho_F}$ is the optimal value of $\mathcal{G}$ when we consider the multimodal fusion-only problem with loss function $L_{\mathcal{G},fusion} = \sum_{\Lambda}^{\{S,F\}} \rho_\Lambda \parallel \mathcal{G} - X_\Lambda W_\Lambda \parallel_F^2$. Intuitively, the prediction term of the overall loss function $K_{pred} \parallel Y - \mathcal{G}\beta \parallel_2^2$ would also impact the optimal value of $\mathcal{G}$. Therefore, instead of using

$\frac{\rho_S X_S W_S + \rho_F X_F W_F}{\rho_S + \rho_F}$ as an estimate of $\mathcal{G}$, we need to find a formula that can directly evaluate the target value on new data using $X_S$, $X_F$, $W_S$, $W_F$, and $\beta$.

To achieve this, we use the fact that $\tilde{Y} \approx Y$ at convergence for the training set, where $\tilde{Y}$ represents the estimated target and $Y$ represents the true target value. Therefore, we have $\tilde{Y} \approx \mathcal{G}\beta$ at convergence, where $\mathcal{G}$ is the optimal latent space with the expression of $\mathcal{G} = (\rho_S X_S W_S + \rho_F X_F W_F + K_{pred} Y \beta^T)(\frac{I}{\rho_S + \rho_F} - \frac{K_{pred}\beta\beta^T}{(\rho_S+\rho_F)^2 + (\rho_S+\rho_F)K_{pred}\beta^T\beta})$. Substituting $Y$ with $\tilde{Y}$ and $\mathcal{G}$ with the above equation, we can get an equation for $\tilde{Y}$ which is only parameterized by $X_S$, $X_F$, $W_S$, $W_F$, and $\beta$:

$$\tilde{Y} = (\rho_S X_S W_S + \rho_F X_F W_F + K_{pred}\,\tilde{Y}\beta^T)(\frac{\beta}{\rho_S+\rho_F} - \frac{K_{pred}\beta\beta^T\beta}{(\rho_S+\rho_F)^2 + (\rho_S+\rho_F)K_{pred}\beta^T\beta})$$

Solving this equation for $\tilde{Y}$ yields $\tilde{Y} = \frac{\rho_S X_S W_S + \rho_F X_F W_F}{\rho_S + \rho_F}\beta$. Notably, the derivation of this result is only based on the assumption that $\tilde{Y} \approx Y$ at convergence for the training set. It does not rely on the equality between $\mathcal{G}$ and $\frac{\rho_S X_S W_S + \rho_F X_F W_F}{\rho_S + \rho_F}$.

Finally, substituting $Y$ with $\frac{\rho_S X_S W_S + \rho_F X_F W_F}{\rho_S + \rho_F}\beta$, we can get (at convergence):

$$\mathcal{G} = (\rho_S X_S W_S + \rho_F X_F W_F + K_{pred}\frac{\rho_S X_S W_S + \rho_F X_F W_F}{\rho_S + \rho_F}\beta\beta^T)(\frac{I}{\rho_S + \rho_F} - \frac{K_{pred}\beta\beta^T}{(\rho_S + \rho_F)^2 + (\rho_S + \rho_F)K_{pred}\beta^T\beta})$$

By simplifying the equation, we can get $\mathcal{G} = \frac{\rho_S X_S W_S + \rho_F X_F W_F}{\rho_S + \rho_F}$. Reiteratively, this result assumes $\tilde{Y} \approx Y$, thus is only valid at convergence.

One question that readers may have is: since $\frac{\rho_S X_S W_S + \rho_F X_F W_F}{\rho_S + \rho_F}$ is the optimal $\mathcal{G}$ value for the multimodal fusion-only problem $L_{\mathcal{G},fusion} = \sum_{\Lambda}^{\{S,F\}} \rho_\Lambda \parallel \mathcal{G} - X_\Lambda W_\Lambda \parallel_F^2$, why the optimal value of $\mathcal{G}$ looks the same after the incorporation of the prediction loss — where does the information of prediction task go? Essentially, although we still write the optimal $\mathcal{G}$ as $\frac{\rho_S X_S W_S + \rho_F X_F W_F}{\rho_S + \rho_F}$, $W_S$ and $W_F$ are now functions of $\beta$ after the incorporation of prediction task: $W_S = W_S(\beta; \lambda_{fusion}, \lambda_{pred}, K_{pred})$, $W_F = W_F(\beta; \lambda_{fusion}, \lambda_{pred}, K_{pred})$. At convergence, $W_S$ and $W_F$ have integrated the information from prediction task in an appropriate way, such that $\mathcal{G} = \frac{\rho_S X_S W_S + \rho_F X_F W_F}{\rho_S + \rho_F}$.

### Reference


1. Savarese, P., Silva, H. & Maire, M. Winning the lottery with continuous sparsification. *Advances in neural information processing systems* **33**, 11380-11390 (2020).
