## Appendix B for "Optimizing Antidepressant Efficacy: Generalizable Multimodal Neuroimaging Biomarkers for Prediction of Treatment Response"

### Appendix B — Significant Characteristics of Latent Space in TOMMF

In this appendix, we discuss the theoretical foundation of two significant characteristics of latent space in the TOMMF framework: 1) data driven dimensionality, and 2) pseudo-exclusive compositions. We also elaborate how these properties are uniquely endowed by the L0-regularization with theoretical analyses and experimental results.

#### *Data Driven Dimensionality — Make Adequate Contribution, or Be Nullified*

One of the significant characteristics of the TOMMF is the data driven dimensionality of latent features. To elaborate this property, let us first revisit the loss function of TOMMF framework:

$$L = \sum_{\Lambda}^{\{S,F\}} (\rho_\Lambda \parallel \mathcal{G} - X_\Lambda W_\Lambda \parallel_F^2 + \lambda_{fusion} \sum_{j=1}^{P} \parallel W_{\Lambda,j} \parallel_0) + K_{pred} \parallel Y - \mathcal{G}\beta \parallel_2^2 + \lambda_{pred} \parallel \beta \parallel_0$$

In this analysis, we examine each dimension $\mathcal{G}_j$ in the latent space $\mathcal{G}$, where $\mathcal{G} = [\mathcal{G}_1, \ldots, \mathcal{G}_j, \ldots, \mathcal{G}_P]$ and $\mathcal{G}_j$'s are column vectors. The predictive weights $\beta$ can also be denoted as $\beta = [\beta_1, \ldots, \beta_j, \ldots, \beta_P]^\top$, where $\beta_j$ is the weight of $\mathcal{G}_j$. We note that for a latent dimension $\mathcal{G}_j$ that exhibits zero correlation with the target $Y$, $\beta_j = 0$ is the solution to the loss function. As a result, $W_{\Lambda,j}$ would also converge to the zero vector to minimize L0-norm penalty. Notably, although we describe the process in a sequential manner, the shrinkage of $\beta_j$ and $W_{\Lambda,j}$ actually happens simultaneously. Hence, we have showed that $\mathcal{G}_j$ would be automatically set to zero if it does not provide contribution to the prediction task. In reality, a non-zero latent dimension $\mathcal{G}_j$ needs to make adequate contribution to the prediction task (the decrease in $K_{pred} \parallel Y - \mathcal{G}\beta \parallel_2^2$) to outweigh the penalty it introduces. In this way, the TOMMF framework achieves a data-driven way to determine the dimensionality of latent features — a new dimension would be included if it benefits the prediction task, and no additional dimensions would be added if the gain in performance is too marginal to outweigh the penalty (Extended Data Figure 1a). Compared to the hard-thresholding, TOMMF does not require an empirical percentage to determine how many features can be incorporated in the model, thus preventing potential issue of over-inclusion and over-exclusion. The sparsity parameters $\lambda_{fusion}$ and $\lambda_{pred}$ in the TOMMF framework only serve as parameters controlling the model complexity — the framework itself judges whether a feature is informative or not.

#### *Pseudo-exclusive Composition — Dimensions Compete for Features*

Another significant characteristic of the TOMMF framework is the pseudo-exclusive composition of latent features. Compared with its L1-regularization counterpart, the framework imposes extra penalty on the occurrence of one feature in multiple dimensions. Specifically, L1-regularization does not impose additional penalty on the loss function as long as the summation of weights across dimensions for a particular feature remains same, while L0-regularization penalizes each occurrence of feature in the latent dimensions. When tuning the sparsity parameter for L0- and L1-regularization, we can observe a significantly different behaviors of these two penalty types— the L0-regularized framework gradually achieves a high level of orthogonality across latent features (Extended Data Figure 1a), whereas the L1-regularized framework gradually aggregate the latent space into one major dimension with a few auxiliary dimensions (Extended Data Figure

1b). The behavior of L1-regularized latent space also aligns with its mathematical foundation — putting everything into one would minimize the penalty on $\beta$. Again, for the L0-regularized framework, the penalty on $\beta$ is also for the non-zero occurrence of its elements, not their amplitudes. As the L1-regularized framework may introduce a few auxiliary dimensions to improve the prediction performance while keeping the penalty low, the L0-regularized framework needs to make every dimension counts. Actually, the L0-regularization enables each feature to make optimal contribution to the dimension it constitutes, and each dimension to make optimal contribution to the prediction target. In this way, the framework offers a data-driven way for decomposing significant dimensions in the overall predictive pattern, allowing us to dissect the intricate psychopharmacology of MDD.
